# Efficacy of an early first dose of whole-cell pertussis vaccine in infants for the prevention of IgE-mediated food allergy: A randomised controlled trial

**DOI:** 10.64898/2026.11.13.26367163

**Authors:** Gladymar Pérez Chacón, James Totterdell, Marie J. Estcourt, Kirsten P. Perrett, Dianne E. Campbell, Julie A. Marsh, Nicholas Wood, Adam W. Bartlett, Phoebe C.M. Williams, Michael D. O’Sullivan, Claire S. Waddington, Nicole L. Messina, Brynn Wainstein, Peter Hsu, Sonia McAlister, Mark A. Jones, Catherine Hughes, Ushma Wadia, Peter B. McIntyre, Nigel Curtis, Michael Gold, Patrick G. Holt, Peter C. Richmond, Tom Snelling

**Author notes:** Corresponding author: Professor Tom Snelling. Head - Health and Clinical Analytics Lab. Sydney School of Public Health, The University of Sydney. Sydney, New South Wales, Australia. Contributed equally (Gladymar Pérez Chacón, James Totterdell).

## Abstract

**Background:** Evidence from mechanistic and observational studies suggests that replacing the first acellular pertussis (aP) vaccine dose in early infancy with a whole-cell pertussis (wP)-containing vaccine may reduce the risk of developing IgE-mediated food allergy during childhood.

**Objective:** We aimed to determine if a dose of wP at approximately 6 weeks old, in lieu of aP vaccine, could reduce the incidence of food allergy in infants.

**Methods:** In this multicentre, randomised, participant and outcome assessor-blinded, parallel, two-stage, Bayesian adaptive trial we assigned Australian infants to one of two pertussis vaccination schedules. These consisted of either a standard homologous priming with a hexavalent combination aP vaccine (diphtheria, tetanus, aP, hepatitis B, inactivated poliovirus types 1, 2, and 3, and *Haemophilus influenzae* type b; DTaP-HepB-IPV-Hib) at approximately 6 weeks, 4 months and 6 months old (‘aP-only’ schedule), or a first dose of a WHO-prequalified pentavalent wP combination vaccine (DTwP-HepB-Hib; Pentabio®, PT Bio Farma Indonesia) at approximately 6 weeks old, followed by hexavalent aP combination vaccine at approximately 4 months and 6 months old (‘mixed wP/aP’ schedule). The primary outcome was physician-confirmed food allergy by age 12 months requiring both evidence of sensitisation and compatible symptoms in response to either incidental or medically supervised oral food challenge. The initial target sample size was 3,000 enrolments to provide approximately 85% power to detect a policy-relevant 3 percentage point reduction in the risk of the primary outcome.

**Results:** A total of 972 infants were enrolled, 486 in each study arm. Of these, 26 of 486 (5.3%) assigned to the aP-only group and 31 of 486 (6.4%) assigned to the mixed wP/aP schedule met the primary endpoint definition. The posterior median difference (95% credible interval) in food allergy among recipients of the mixed wP/aP schedule compared to recipients of the aP-only schedule was 0.01 (−0.02, 0.04; probability [<0] 0.25). There were six serious adverse events (SAEs) among children receiving the aP-only group versus 12 among children receiving the mixed wP/aP schedule; one SAE (hospitalisation for irritability; length of stay < 48 hours) was deemed related to wP vaccine. The trial was stopped early owing to operational reasons.

**Conclusions:** There is a low probability of a policy-relevant reduction in food allergy in infants receiving mixed wP/aP versus aP-only pertussis vaccine schedules. Funded by the National Health and Medical Research Council of Australia; ANZCTR number, ACTRN12617000065392p.

**Clinical implications:** An early first dose of wP is unlikely to afford a policy-relevant reduction in the probability of developing food allergy, eczema or atopic sensitisation in infants.

**Capsule summary:** An early first dose of a 5-in-1 wP combination vaccine is reasonably tolerated but would not be expected to have a material impact on the prevalence of food allergy in infants.

## Introduction

IgE-mediated food allergy affects approximately 10% of Australian infants and is a leading cause of anaphylaxis presentations in children, which have increased substantially in the last 25 years.[1,2] Since 2016, national guidelines have promoted the introduction of solid foods between the ages of 4 and 6 months old.[3] However, compared to the effect of delayed introduction of solids found in clinical trials, the population-level impact of these guidelines on rates of food allergy has been more modest.[4–6]

Food allergies are thought to arise in young children whose immune systems fail to mature into a balanced Th_1_/Th_17_/Th_2_ phenotype, with persistence of a Th_2_-skewed phenotype characterised by production of IL-4, IL-5 and IL-13.[7] The latter promotes class-switching to food allergen-specific IgE. Some exposures in early life, like vaccination, might modulate the maturation of immune responses and have emerged as new potential targets for preventing food allergy.[8]

The whole-cell pertussis (wP) vaccine has been a foundational component of the global Expanded Program of Immunisation since its inception in 1974, and has been administered to at least 1.4 billion children in Gavi-supported countries since 2000.[9–11]. However, adverse events following immunisation and heterogeneity in wP vaccine manufacturing processes have dominated considerations in pertussis vaccine policy in recent decades.[10,12,13]

Despite its high effectiveness in preventing pertussis notifications, Australia progressively withdrew a locally manufactured wP-based combination vaccine from its National Immunisation Program (NIP) between 1997 and 1999 due to reactogenicity concerns, and replaced it with an aP-based combination vaccine.[14,15] A three-dose aP-only primary schedule has been used in Australia since the late 1990s, and this schedule is now recommended in most high income countries. In these settings, aP booster doses and/or maternal pertussis immunisation have been implemented to mitigate waning immunity and protect infants who are too young to be vaccinated.[16,17] Conversely, vaccination with wP-based formulations is the primary strategy for preventing severe pertussis disease in infancy in most low- and middle-income countries.[10]

wP vaccines induce a Th_1_ and Th_17_-polarised immunophenotype which persists into early adulthood,[18] an effect that might be driven by the adjuvant properties of lipopolysaccharide within the whole-killed *Bordetella pertussis* cells.[19] Compared to aP vaccines, wP vaccines have been reported to attenuate IgE-mediated immune responses to diphtheria (DT) and pertussis toxoids (PT) vaccine antigens,[20–22] although data on the effects of wP vaccines on tetanus toxoid (TT) have been conflicting.[23,24]

We examined Australian children born during the transition from wP to aP vaccines in a case-control study; those who developed IgE-mediated food allergy were less likely to have received wP than aP as their first pertussis vaccine dose in early infancy (odds ratio [OR] 0.77; 95% confidence interval [CI], 0.62–0.95).[25]

Given the apparent in vitro effects of wP versus aP priming on immunophenotype and the association with food allergy we observed in our case-control study, we hypothesised that compared to a standard aP-only vaccine schedule, substituting the first aP vaccine dose with wP would help induce a balanced Th_1_/Th_17_/Th_2_ immune profile and thereby prevent the development of food allergy in infants.[26]

## Methods

### Trial design and participants

OPTIMUM was a two-stage, participant and outcome assessor-blinded, Bayesian group sequential randomised trial, conducted at four study sites: Perth Children’s Hospital (Western Australia), Murdoch Children’s Research Institute (Victoria), Sydney Children’s Hospital, Randwick and The Children’s Hospital at Westmead (New South Wales).[26,27]

We enrolled healthy infants born on or after 32 weeks gestation. They were randomised 1:1 to receive either a standard three-dose homologous pertussis priming schedule with a 6-in-1 aP combination vaccine at approximately 6 weeks, 4 months and 6 months old (‘aP-only schedule’) or a heterologous pertussis priming schedule consisting of a first dose of a WHO-prequalified 5-in-1 wP combination vaccine at 6 weeks old followed by the 6-in-1 aP combination vaccine at 4 months and 6 months old (‘mixed wP/aP schedule’).

The aP combination vaccine (DTaP_3_-HepB-IPV-Hib; Infanrix hexa®, GlaxoSmithKline Australia; 0.5 mL) contained three proteins purified from cultures of *B pertussis* (pertussis toxin, filamentous haemagglutinin and pertactin), diphtheria toxoid, tetanus toxoid, recombinant hepatitis B surface antigen, inactivated poliovirus types 1, 2, and 3, and *Haemophilus influenzae* type b capsular polysaccharide conjugated to tetanus toxoid. In July 2023, an alternative 6-in-1 aP combination vaccine containing pertussis toxin, filamentous haemagglutinin, pertactin and fimbriae types 2 and 3 was added to the NIP (DTaP_5_-HepB-IPV-Hib capsular polysaccharide conjugated to outer membrane protein complex of *Neisseria meningitidis*; Vaxelis®, Maxx Pharma Pty Ltd; 0.5 mL). The wP combination vaccine (DTwP-HepB-Hib; Pentabio®, PT Bio Farma, Indonesia; 0.5 mL) contained diphtheria toxoid, tetanus toxoid, inactivated whole-cell *B pertussis*, recombinant hepatitis B surface antigen, and *H influenzae* type b capsular polysaccharide conjugated to tetanus toxoid.[28]

To ensure each child received at least three primary doses of IPV while maintaining blinding, DTaP-IPV (Infanrix IPV, GlaxoSmithKline Australia; 0.5 mL intramuscular) was administered to all infants at 18 months old, in lieu of a dose of DTaP (without IPV) per the Australian NIP. Infants with planned travel to a polio-affected region (as defined by the Global Polio Eradication Initiative) at any time before the completion of the study were ineligible at enrolment,[29] with supplementary IPV vaccination facilitated for any unplanned travel to increased risk countries before 18 months. The wP combination vaccine (either single dose or multidose vials) was procured directly from its manufacturer (PT Bio Farma, Indonesia) for the implementation of this trial; all other vaccines were sourced from Australia’s NIP.

Stage one was designed to compare IgE-mediated immune responses to TT post-priming and boosting in each group and involved the first 150 infants enrolled at Perth Children’s Hospital only.[27] A comparison of anti-TT IgE responses and solicited adverse reactions at 6 and 7 months old, and IgG responses to vaccine antigens at 6, 7, 18, and 19 months old between the study arms has been reported previously.[23,30]

Stage two involved all four study sites with more streamlined follow-up. A single phlebotomy visit was scheduled for the first 150 infants enrolled at Perth Children’s Hospital only. In this subset of infants, we also solicited adverse event data following the first pertussis vaccine dose at age six weeks old and the first booster dose at age 18 months old.

### Trial procedures

The study protocol and statistical analysis plan have been published previously and are available through open access.[26,27] Briefly, participants were enrolled between 6 and < 12 weeks old and were randomised with equal probability to the aP-only schedule versus the mixed wP/aP schedule. The allocation sequence was generated by the trial statistician using computer block randomisation with block sizes of six, eight, and 10, stratified by study site and concealed in opaque envelopes.

At approximately 6 weeks old, the randomly assigned pertussis containing vaccine was administered intramuscularly into the anterolateral aspect of the right thigh by an unblinded research nurse who concealed the allocation and who had no further involvement in study visits or trial procedures. A 15 mg/kg dose of oral paracetamol was given to all infants before the vaccine was administered; parents were recommended to administer two further paracetamol doses six hours apart, but this was not mandated. The NIP-scheduled 13-valent pneumococcal conjugate vaccine and monovalent rotavirus vaccine (RV1) were coadministered.[30]

At the discretion of each study site, infants enrolled in stage two underwent a baseline and follow-up SCORAD (SCORing Atopic Dermatitis) assessments or alternative clinical reporting methods, at enrolment and 12 months old to measure the extent and severity of any skin inflammation.[31] Parents were provided and encouraged to follow the Australasian Society of Allergy and Clinical Immunology infant feeding guidelines for allergy prevention.[3] The guidelines recommend the introduction of solids, including peanuts and cooked hen’s egg, once the infant is developmentally ready; this is generally at around 6 months old and not before 4 months old. Enrolled infants were also recommended complementary feeding with other common food allergens by 12 months old, with emphasis on foods regularly consumed by the family.[3] Either the age (stage one) or date (stage two) of introduction of potentially allergenic solid foods was reported at each scheduled study visit and via phone or electronic contact. A skin prick test (SPT) was scheduled for all infants at approximately 12 months old and no later than 19 months. Protocol-specified allergens (Immunotek, Madrid, Spain) were cow’s milk, hen’s egg, peanut, cashew, house dust mite, cat, and rye grass.[26] At the discretion of each site principal investigator, additional unscheduled SPTs were permitted as part of the routine assessment of infants with suspected IgE-mediated food reactions. Sensitisation to food allergens or a positive SPT for a specific food was defined as an SPT wheal size > 1 mm greater than that produced by a negative control (saline buffer/50% glycerol), in the context of a positive histamine control and a negative saline control. Children with a positive SPT underwent a blinded clinical assessment by the lead allergist at each site for suitability for an oral food challenge (OFC, Supplementary material 2, Figure E1) from 12 months old (and before 19 months). At their discretion, children underwent an OFC unless there was a clear history of tolerance or an unequivocal history of anaphylaxis to the food. OFCs followed site-specific procedures and ASCIA Food Allergen Challenge Protocol Dosage Guide (Supplementary material 1).[32–36] Briefly, an initial dose of 1 to 10 mg of food protein was administered, followed by additional doses that increased semi-logarithmically or by doubling of the protein quantity. OFCs were scored by an investigator blinded to the group allocation using the PRACTALL scoring system.[37]

At 19 months old, total IgE, TT, egg white, whole egg, peanut, cashew, cow’s milk, *Dermatophagoides pteronyssinus*, cat, and rye grass-specific IgE concentrations were measured in plasma using the ImmunoCAP assay (Thermo Fisher Scientific, Uppsala, Sweden).

Safety monitoring, data collection, case definitions and reporting of adverse events (AEs) followed national guidelines.[38] In stage one, we captured unsolicited AEs occurring within 7 days of each pertussis vaccine dose scheduled at 6 weeks, 4 months, 6 months and 18 months old. In stage two, we exclusively captured unsolicited AEs at 6 weeks and 18 months old for the first 150 study participants enrolled at Perth Children’s Hospital. Solicited adverse reactions following the primary pertussis vaccine series in the stage one cohort have been reported previously.[23]

Serious adverse events (SAEs) considered related to either study vaccine administered at 6 weeks old or to any study procedure were solicited throughout the study period. SAEs deemed unrelated to the study vaccines or study procedures were collected from enrolment at approximately 6 weeks old to 6 months post-randomisation. Predefined adverse events of special interest (AESIs), including breakthrough pertussis infection and hypotonic hyporesponsive episodes, were collected throughout the study.[26] All AEs were coded according to the Medical Dictionary for Regulatory Activities (MedDRA) System Organ Class categorisation and were reviewed quarterly by an independent Data Safety and Monitoring Committee (DSMC).[39]

### Trial outcomes

The primary outcome was IgE-mediated food allergy by 12 months old. Participants met the primary endpoint if they met either of the pre-specified definitions for (1) unequivocal or (2) highly probable IgE-mediated food allergy. Unequivocal IgE-mediated food allergy was defined as sensitisation on SPT (>1mm greater than the negative control) to the food of interest, and either: (a) a positive OFC, or (b) a history of clinician-diagnosed food anaphylaxis (symptoms affecting at least two of the following: skin, gastrointestinal tract, respiratory tract, cardiovascular system). Highly probable IgE-mediated food allergy was defined as a history of food allergic reaction consistent with PRACTALL criteria, with evidence of sensitisation on SPT to the food of interest.[37]

Additional clinical and mechanistic (secondary) outcomes were new-onset parent-reported clinician-diagnosed atopic eczema by (1) 6 or (2) 12 months old with a positive SPT to any allergen by 12 months old (onset defined by the documented age at first occurrence), (3) a SPT more than 1 mm greater than the negative control or (4) at least 3 mm greater than the negative control, to at least one allergen by 12 months old.

Outcomes related to IgE-mediated immune responses in plasma included total IgE and allergen-specific IgE (kU/L) at 19 months old as specified above.

### Trial oversight

Approval was obtained from the Child and Adolescent Health Service Human Research Ethics Committee (RGS0000000019). Written informed consent was obtained from a parent or guardian of each enrolled infant. An independent DSMC provided safety oversight. A consumer representative with lived experience of vaccine-preventable disease was a member of the OPTIMUM Trial Steering Committee since before enrolment commenced, and contributed to decisions on trial conduct, the decision to terminate enrolment early, and interpretation of the findings reported here.

The trial was registered with the Australian New Zealand Clinical Trial Registry (ACTRN12617000065392p: https://www.anzctr.org.au/Trial/Registration/TrialReview.aspx?id=371998&isReview=true; December 15, 2016) and funded by Australia’s National Health and Medical Research Council (GNT1158722) and the Telethon New Children’s Hospital Research Fund 2012 (Round 1). The funders had no involvement in the study design, data collection, analysis, interpretation, or writing of the report.

### Statistical analysis

This trial used an adaptive group sequential design and Bayesian inference as detailed in the protocol and the statistical analysis plan.[27] Analyses were performed according to intent-to-treat with participants grouped per their randomly assigned schedule. For all summaries and pre-specified analyses, all available outcomes were included.

The primary analysis used independent beta-binomial models to estimate the difference in probability of IgE-mediated food allergy between the two vaccine strategies; uniform priors were assumed for both response probabilities. Secondary analyses of the primary outcome assumed a logistic regression model for occurrence of IgE-mediated food allergy, adjusting for study site, sex, breastfeeding status, birth order, parental income, mode of delivery, and family history of atopic disease at baseline. Participants with no SPT result were assumed to not meet the primary outcome. A sensitivity analysis excluded those with no SPT result.

A posterior probability of any reduction in IgE-mediated food allergy of at least 0.95 from the primary analysis was considered evidence of superiority of the mixed wP/aP over the aP-only schedule. A first interim analysis was scheduled to occur after the 200th participant completed follow-up. Additional interim analyses were to occur for every additional 200 participants unless a stopping rule was met or the maximum sample size had been reached. At each interim analysis, the trial could be stopped for expected success or futility if the predicted probability of meeting the superiority condition was >0.95 or <0.05, respectively, based on the posterior predictive distribution from the primary analysis model.[27]

The stopping thresholds above were chosen based on trial simulations to estimate their effects on trial operating characteristics across various enrolment rates. Specifically, we assessed type I error, expected stopping times, and power to detect a 3-percentage point reduction in food allergy which we considered by the investigators to be the smallest effect that would be relevant to policy. Under the specified thresholds and assuming 10% incidence of IgE-mediated food allergy by age 12 months in the aP-only group, simulations indicated that enrolling 3,000 infants would ensure 85% power for detecting a 3-percentage point reduction (to 7%) in the mixed wP/aP group. Before any efficacy analyses were conducted, the maximum sample size was revised to 2,000 participants due to slow enrolment and resource constraints. Statistical power was reassessed based on the same thresholds indicating the revised maximum sample size afforded 73% power to detect a 3-percentage point reduction in food allergy (from 10% to 7%).

Analyses of the clinical and mechanistic secondary outcomes followed the same approach as the primary outcome. Analyses of total and specific IgE concentrations at 19 months assumed unadjusted log-normal linear regression models to estimate geometric mean ratios, with IgE concentrations reported as <0.10 kU/L were treated as left-censored at 0.005 kU/L, and logistic regression models to estimate differences in the probability of exceeding the prespecified thresholds of 0.01 kU/L and 0.1 kU/L.

Analyses were performed in R version 4.6.1 using Stan version 2.39.0, via CmdStanR version 0.9.0.[40–42]

## Results

### Trial timeline

Enrolment in stage one commenced at Perth Children’s Hospital in March 2018 and follow-up was completed in July 2021. Enrolment in stage two commenced in January 2020. An interim analysis was performed in September 2022 which included 200 participants who had completed the protocol (of 435 participants enrolled by that time); no stopping rule was met. In December 2023, the trial steering committee terminated enrolment prior to any further analyses because we were unable to procure further doses of the wP vaccine approved for this study. By then, 972 infants had been enrolled. Follow-up for the final enrolled participant was completed on June 6th, 2025.

### Participants

In total, parents of 6,056 infants were invited to participate, of whom 975 were assessed in clinic for trial eligibility. Of the 972 subsequently randomised, 486 were assigned to each study arm (Figure 1, Table 1, Supplementary material 2, Table E1). All except for one infant received either a three-component aP (Infanrix hexa) or wP (Pentabio) vaccine for their first pertussis vaccine dose per their random assignment. All were included in the analysis of the primary outcome (Table 2).

**Figure 1:**
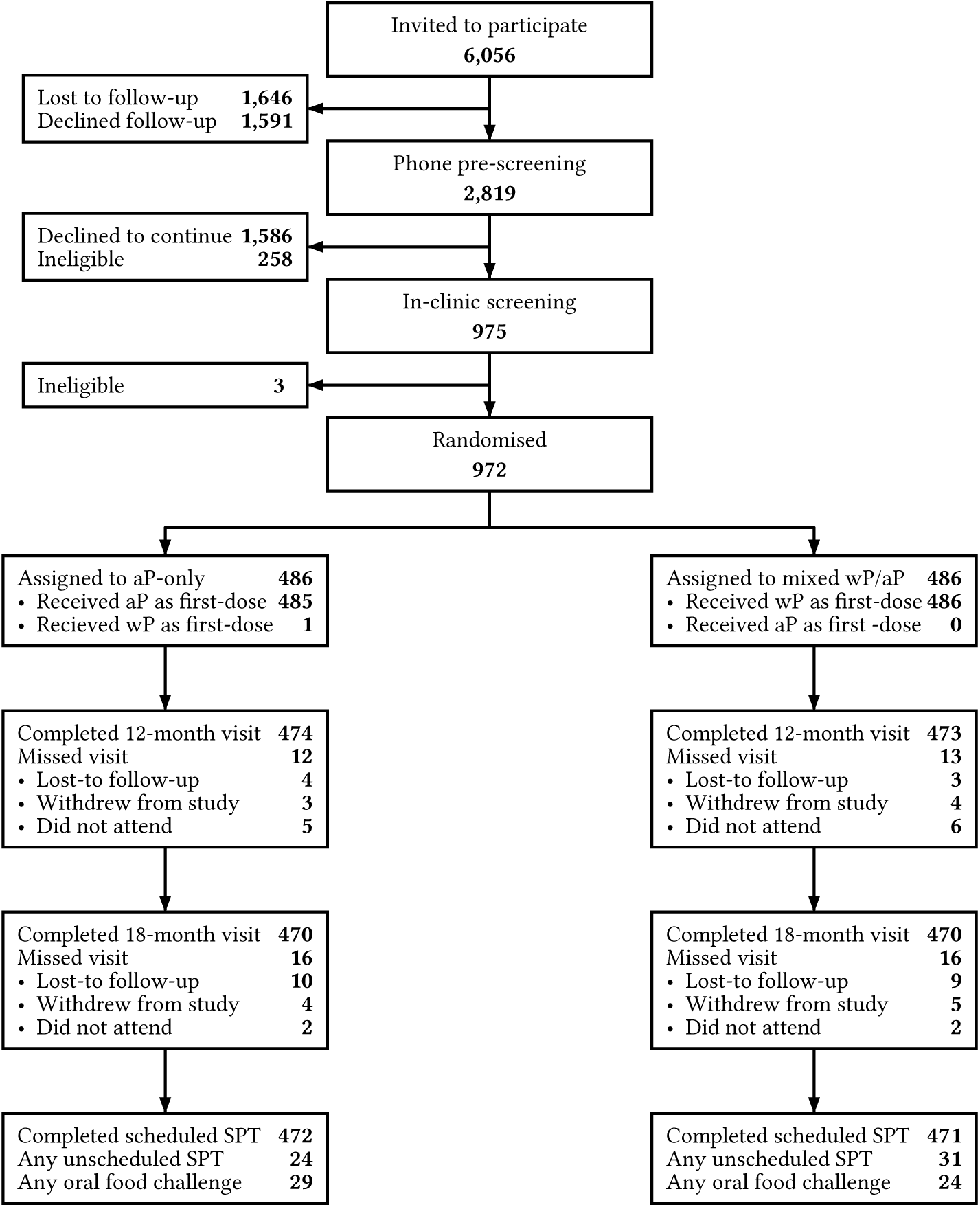
Trial flowchart

**Table 1:** Baseline characteristics of trial participants.

| **Characteristic** | **N** | **aP**<br>N = 486<sup>1</sup> | **wP**<br>N = 486<sup>1</sup> | **Overall**<br>N = 972<sup>1</sup> |
|---|---|---|---|---|
| **Protocol stage** | 972 |  |  |  |
| 1 |  | 75 (15%) | 75 (15%) | 150 (15%) |
| 2 |  | 411 (85%) | 411 (85%) | 822 (85%) |
| **Participating site** | 972 |  |  |  |
| CHW |  | 30 (6%) | 29 (6%) | 59 (6%) |
| MCRI |  | 220 (45%) | 219 (45%) | 439 (45%) |
| PCH |  | 206 (42%) | 207 (43%) | 413 (42%) |
| SCHN |  | 30 (6%) | 31 (6%) | 61 (6%) |
| **Gender** | 972 |  |  |  |
| Female |  | 221 (45%) | 242 (50%) | 463 (48%) |
| Male |  | 265 (55%) | 244 (50%) | 509 (52%) |
| **Age at enrolment (days)** | 972 | 47 (44, 50) | 47 (44, 50) | 47 (44, 50) |
| **Combined parental income status** | 971 |  |  |  |
| < \$18,000 |  | 1 (0%) | 0 (0%) | 1 (0%) |
| \$18,000 to \$37,000 |  | 1 (0%) | 4 (1%) | 5 (1%) |
| \$37,001 to \$87,000 |  | 22 (5%) | 20 (4%) | 42 (4%) |
| \$87,001 to \$180,000 |  | 197 (41%) | 222 (46%) | 419 (43%) |
| >\$180,000 |  | 264 (54%) | 240 (49%) | 504 (52%) |
| (Missing) |  | 1 | 0 | 1 |
| **Birth country of mother** | 959 |  |  |  |
| Australia |  | 372 (78%) | 378 (79%) | 750 (78%) |
| United Kingdom |  | 17 (4%) | 17 (4%) | 34 (4%) |
| New Zealand |  | 14 (3%) | 10 (2%) | 24 (3%) |
| Malaysia |  | 4 (1%) | 13 (3%) | 17 (2%) |
| Ireland |  | 5 (1%) | 6 (1%) | 11 (1%) |
| China (excludes SARs and Taiwan) |  | 4 (1%) | 6 (1%) | 10 (1%) |
| Other |  | 64 (13%) | 49 (10%) | 113 (12%) |
| (Missing) |  | 6 | 7 | 13 |
| **Birth country of father** | 953 |  |  |  |
| Australia |  | 362 (76%) | 350 (74%) | 712 (75%) |
| United Kingdom |  | 28 (6%) | 21 (4%) | 49 (5%) |
| New Zealand |  | 13 (3%) | 20 (4%) | 33 (3%) |
| Ireland |  | 8 (2%) | 10 (2%) | 18 (2%) |
| India |  | 7 (1%) | 4 (1%) | 11 (1%) |
| Other |  | 60 (13%) | 70 (15%) | 130 (14%) |
| (Missing) |  | 8 | 11 | 19 |
| **Mother's ethnicity**<sup>2</sup> | 972 |  |  |  |
| European Caucasian |  | 425 (87%) | 405 (83%) | 830 (85%) |
| Asian |  | 46 (9%) | 68 (14%) | 114 (12%) |
| Indian subcontinent |  | 10 (2%) | 17 (3%) | 27 (3%) |
| South American |  | 7 (1%) | 4 (1%) | 11 (1%) |
| Indigenous Australia |  | 6 (1%) | 1 (0%) | 7 (1%) |
| Pacific Islander |  | 2 (0%) | 4 (1%) | 6 (1%) |
| Black African |  | 0 (0%) | 2 (0%) | 2 (0%) |
| **Father's ethnicity**<sup>2</sup> | 972 |  |  |  |
| European Caucasian |  | 433 (89%) | 414 (85%) | 847 (87%) |
| Asian |  | 34 (7%) | 44 (9%) | 78 (8%) |
| Indian subcontinent |  | 13 (3%) | 15 (3%) | 28 (3%) |
| Pacific Islander |  | 6 (1%) | 11 (2%) | 17 (2%) |
| South American |  | 7 (1%) | 4 (1%) | 11 (1%) |
| Indigenous Australia |  | 3 (1%) | 3 (1%) | 6 (1%) |
| Black African |  | 1 (0%) | 3 (1%) | 4 (0%) |
| **Infant's ethnicity**<sup>2</sup> | 972 |  |  |  |
| European Caucasian |  | 449 (92%) | 442 (91%) | 891 (92%) |
| Asian |  | 51 (10%) | 76 (16%) | 127 (13%) |
| Indian subcontinent |  | 17 (3%) | 21 (4%) | 38 (4%) |
| South American |  | 11 (2%) | 6 (1%) | 17 (2%) |
| Pacific Islander |  | 4 (1%) | 10 (2%) | 14 (1%) |
| Indigenous Australia |  | 8 (2%) | 5 (1%) | 13 (1%) |
| Black African |  | 1 (0%) | 4 (1%) | 5 (1%) |
| **First born** | 972 | 278 (57%) | 273 (56%) | 551 (57%) |
| **Feeding status** | 972 |  |  |  |
| Both breastfed and formula-fed |  | 111 (23%) | 140 (29%) | 251 (26%) |
| Both breastfed and started on solids |  | 1 (0%) | 0 (0%) | 1 (0%) |
| Exclusively breastfed |  | 343 (71%) | 321 (66%) | 664 (68%) |
| Exclusively formula-fed |  | 31 (6%) | 25 (5%) | 56 (6%) |
| **Cat ownership** | 972 |  |  |  |
| No |  | 373 (77%) | 374 (77%) | 747 (77%) |
| Inside |  | 57 (12%) | 62 (13%) | 119 (12%) |
| Outside |  | 2 (0%) | 3 (1%) | 5 (1%) |
| Both |  | 54 (11%) | 47 (10%) | 101 (10%) |
| **Dog ownership** | 971 |  |  |  |
| No |  | 239 (49%) | 278 (57%) | 517 (53%) |
| Inside |  | 57 (12%) | 48 (10%) | 105 (11%) |
| Outside |  | 12 (2%) | 10 (2%) | 22 (2%) |
| Both |  | 177 (36%) | 150 (31%) | 327 (34%) |
| (Missing) |  | 1 | 0 | 1 |
| **Maternal gravidity** | 971 |  |  |  |
| 1 |  | 213 (44%) | 205 (42%) | 418 (43%) |
| 2 |  | 144 (30%) | 154 (32%) | 298 (31%) |
| 3 |  | 69 (14%) | 70 (14%) | 139 (14%) |
| 4 |  | 24 (5%) | 32 (7%) | 56 (6%) |
| 5 |  | 16 (3%) | 5 (1%) | 21 (2%) |
| 6+ |  | 19 (4%) | 20 (4%) | 39 (4%) |
| (Missing) |  | 1 | 0 | 1 |
| **Maternal parity** | 971 |  |  |  |
| 1 |  | 284 (59%) | 279 (57%) | 563 (58%) |
| 2 |  | 150 (31%) | 166 (34%) | 316 (33%) |
| 3 |  | 41 (8%) | 30 (6%) | 71 (7%) |
| 4+ |  | 10 (2%) | 11 (2%) | 21 (2%) |
| (Missing) |  | 1 | 0 | 1 |
| **Delivery type** | 971 |  |  |  |
| Elective caesarean section |  | 116 (24%) | 121 (25%) | 237 (24%) |
| Emergency caesarean section |  | 97 (20%) | 108 (22%) | 205 (21%) |
| Forceps/vacuum assisted delivery |  | 69 (14%) | 67 (14%) | 136 (14%) |
| Vaginal delivery |  | 204 (42%) | 189 (39%) | 393 (40%) |
| (Missing) |  | 0 | 1 | 1 |
| **Intrapartum Antibiotics?** | 962 | 166 (35%) | 146 (30%) | 312 (32%) |
| (Missing) |  | 6 | 4 | 10 |
| **Neonatal systemic antibiotics (first 28 days of life)?** | 971 | 41 (8%) | 35 (7%) | 76 (8%) |
| (Missing) |  | 1 | 0 | 1 |
| **Gestational age at delivery (weeks)** | 972 | 39 (38, 40) | 39 (38, 39) | 39 (38, 39) |
| **Weight (grams)** | 970 | 3,420 (3,140, 3,750) | 3,400 (3,100, 3,760) | 3,405 (3,120, 3,752) |
| (Missing) |  | 2 | 0 | 2 |
| **Length (cm)** | 965 | 51 (49, 52) | 50 (49, 52) | 50 (49, 52) |
| (Missing) |  | 4 | 3 | 7 |
| **Head Circumference (cm)** | 963 | 35 (34, 36) | 35 (34, 36) | 35 (34, 36) |
| (Missing) |  | 5 | 4 | 9 |
| **Apgar score (1 minute)** | 963 |  |  |  |
| 1-7 |  | 56 (12%) | 65 (13%) | 121 (13%) |
| 8 |  | 51 (11%) | 67 (14%) | 118 (12%) |
| 9 |  | 372 (78%) | 351 (73%) | 723 (75%) |
| 10 |  | 1 (0%) | 0 (0%) | 1 (0%) |
| (Missing) |  | 6 | 3 | 9 |
| **Apgar score (5 minutes)** | 963 |  |  |  |
| 1-7 |  | 6 (1%) | 10 (2%) | 16 (2%) |
| 8 |  | 14 (3%) | 20 (4%) | 34 (4%) |
| 9 |  | 434 (90%) | 429 (89%) | 863 (90%) |
| 10 |  | 26 (5%) | 24 (5%) | 50 (5%) |
| (Missing) |  | 6 | 3 | 9 |
| **Maternal pertussis vaccination during preceding pregnancy?** | 971 | 465 (96%) | 459 (95%) | 924 (95%) |
| (Missing) |  | 0 | 1 | 1 |
| **Maternal pertussis Vaccine administered** | 909 |  |  |  |
| Adacel |  | 169 (37%) | 161 (35%) | 330 (36%) |
| Boostrix |  | 238 (52%) | 240 (53%) | 478 (53%) |
| Unknown |  | 48 (11%) | 53 (12%) | 101 (11%) |
| (Missing) |  | 31 | 32 | 63 |
| **Other maternal pertussis vaccination (last 5 years)** | 968 | 240 (50%) | 238 (49%) | 478 (49%) |
| (Missing) |  | 2 | 2 | 4 |
| **Maternal seasonal influenza vaccination during pregnancy?** | 966 | 381 (79%) | 369 (77%) | 750 (78%) |
| (Missing) |  | 1 | 5 | 6 |
| **Family history of atopy** | 972 | 418 (86%) | 415 (85%) | 833 (86%) |

<sup>*1*</sup>n (%); Median (Q1, Q3)
<sup>*2*</sup>Multiple may apply to each participant

**Table 2:** Primary and secondary outcomes.

| **Outcome** | **aP**<br>N = 486 | **wP**<br>N = 486 | **Overall**<br>N = 972 | **%pt. difference**, wP - aP<br>Med. (95% CrI), Pr(<0)<sup>†</sup> |
|---|---|---|---|---|
| **Primary outcome** |  |  |  |  |
| *IgE-mediated food allergy* | 26/486 (5.3%) | 31/486 (6.4%) | 57/972 (5.9%) | 1.0 (-2.0, 4.0), 0.25 |
| Condition 1.a (OFC) | 13 (2.7%) | 9 (1.9%) | 22 (2.3%) |  |
| Condition 1.b (anaphylaxis) | 1 (0.2%) | 1 (0.2%) | 2 (0.2%) |  |
| Condition 2 (highly-probable) | 12 (2.5%) | 21 (4.3%) | 33 (3.4%) |  |
| **Secondary outcomes** |  |  |  |  |
| *Eczema*<sup>‡</sup> |  |  |  |  |
| 6-months | 87/484 (18%) | 78/480 (16%) | 165/964 (17%) |  |
| With SPT sensitisation | 34/483 (7%) | 28/478 (6%) | 62/961 (6%) | -1.2 (-4.3, 2.0), 0.77 |
| 12-months | 145/484 (30%) | 129/480 (27%) | 274/964 (28%) |  |
| With SPT sensitisation | 47/482 (10%) | 43/478 (9%) | 90/960 (9%) | -0.7 (-4.5, 3.0), 0.65 |
| *SPT sensitisation*<sup>§</sup> |  |  |  |  |
| >1mm | 81/473 (17%) | 86/471 (18%) | 167/944 (18%) | 1.1 (-3.7, 6.0), 0.32 |
| ≥3mm | 58/473 (12%) | 61/471 (13%) | 119/944 (13%) | 0.7 (-3.6, 4.9), 0.38 |

<sup>†</sup>%pt. - percentage point, Med - posterior median, CrI - posterior credible interval, Pr(<0) - posterior probability of a reduction. All model estimates are based on the primary analysis Beta-Binomial model.
<sup>‡</sup>There were 8 participants with eczema reported at the time of enrolment, 2 assigned to aP and 6 assigned to wP. These were excluded from new-onset eczema cases.
<sup>§</sup>There were 28 participants with no skin prick test results available before 19-months of age.

Overall, 463 (48%) of 972 infants were female, 924 (95%) were born to a mother who received dTaP in pregnancy, and 833 (86%) had a parent-reported history of food allergy, allergic rhinitis or asthma in at least one first-degree relative. Baseline characteristics were similar across the study groups (Table 1).

There were 930 (96%) infants who completed both study visits at 12- and 18-months old, (466/486 aP-only and 464/486 mixed wP/aP). Scheduled SPTs were performed on time for 943 (98%) participants. Results from additional unscheduled SPTs were available for 55 (5.7%) infants, and 53 (5.5%) infants underwent an OFC (29/486 aP-only and 24/486 mixed wP/aP).

The parent-reported age at introduction of egg, cow’s milk, peanut, fruit, and vegetables were similar across the study groups (Supplementary material 2, Figure E2). At 12-months old, 866/955 (91%) children were currently eating eggs, 792/955 (83%) cow’s milk, and 865/955 (91%) peanut or other nuts; the proportions were similar across the study groups (Supplementary material 2, Table E2).

### Food allergy

The primary endpoint of unequivocal or highly likely food allergy was met in 26 of 486 (5.3%) in the aP-only group and 31 of 486 (6.4%) in the mixed wP/aP group (Table 2). This included 13 of 486 (2.7%) in the aP-only group and 9 of 486 (1.9%) in the mixed wP/aP group with OFC-proven food allergy. The foods most commonly implicated were egg [39 of 972 (4.0%)], peanut [18 of 972 (1.9%)], cow’s milk [16 of 972 (1.6%)], and cashew [5 of 972 (0.5%)] which were similar across the study groups (Table 3).

**Table 3:** Number of infants who met the primary outcome criteria by the reported food allergy. More than one may apply to each infant.

| **IgE-mediated food allergy** | **aP**<br>N = 486 | **wP**<br>N = 486 | **Overall**<br>N = 972 |
|---|---|---|---|
| At least one food allergy | 26 (5.3%) | 31 (6.4%) | 57 (5.9%) |
| Egg | 20 (4.1%) | 19 (3.9%) | 39 (4.0%) |
| Peanut | 6 (1.2%) | 12 (2.5%) | 18 (1.9%) |
| Cow’s milk | 8 (1.6%) | 8 (1.6%) | 16 (1.6%) |
| Cashew | 2 (0.4%) | 3 (0.6%) | 5 (0.5%) |
| Almond | 1 (0.2%) | 0 (0.0%) | 1 (0.1%) |
| Beef | 1 (0.2%) | 0 (0.0%) | 1 (0.1%) |
| Coconut | 1 (0.2%) | 0 (0.0%) | 1 (0.1%) |
| Hazelnut | 0 (0.0%) | 1 (0.2%) | 1 (0.1%) |
| Pecan | 1 (0.2%) | 0 (0.0%) | 1 (0.1%) |
| Sesame | 1 (0.2%) | 0 (0.0%) | 1 (0.1%) |
| Soy | 1 (0.2%) | 0 (0.0%) | 1 (0.1%) |

The incidence of food allergy was 1.0 percentage point higher in the mixed wP/aP group than in the aP-only group (95% CrI: −2.0, 4.0); the probability of any reduction in IgE-mediated food allergy under the mixed wP/aP was 0.25. Covariate adjustment and excluding children without an SPT result had little effect on the estimate (Supplementary material 2, Tables E3 and E4; Figures E3 and E5). The incidence of OFC-proven food allergy was 0.8 percentage points lower in the mixed wP/aP group than in the aP-only group (95% CrI −2.8, 1.1); the probability of any reduction was 0.80.

Post hoc analyses found no evidence of any important subgroup-specific effects by season of birth, ethnicity or family history of atopy (Supplementary material 2, Figure E4). At the time of termination, the predicted probability of eventually meeting our pre-specified criterion for trial success (i.e. if we had not been forced to stop early) was <0.01.

### New-onset eczema

New-onset parent-reported, clinician-diagnosed eczema occurred in 165 of 964 (18%) of infants by 6 months and 274 of 964 (28%) by 12 months old (Table 2). The incidence of eczema was 1.2 (95% CrI −4.3, 2.0) and 0.7 (95% CrI −4.5, 3.0) percentage points lower in the mixed wP/aP group than in the aP-only group by 6 months and 12 months old respectively, and the probability of any reduction was 0.77 and 0.65. Results from the covariate-adjusted analysis were similar (Supplementary material 2, Table E5 and Figure E6).

### Skin prick testing

Most participants had at least one SPT result (944 of 972, 97%). Of these 81/473 (17%) in the aP-only group and 86/471 (18%) in the mixed wP/aP group had evidence of SPT sensitisation (1 mm > negative control; Table 4). The most common allergens resulting in SPT sensitisation were egg [94/942 (10%)], peanut [41/939 (4%)], and cow’s milk [35/940 (4%)] with similar rates across the study groups (Table 4). The difference in SPT sensitisation between the mixed wP/aP group and the aP-only group was 1.1 (95% CrI −3.7, 6.0) percentage points with a probability of reduction in SPT sensitisation of 0.32. The results from the covariate-adjusted analyses were similar (Supplementary material 2, Table E6 and Figure E7). Among infants with any SPT reaction (wheal size >0mm), the distribution of wheal diameters was similar between the study groups (Supplementary material 2, Figure E8).

**Table 4:** Number and percentage of infants with any SPT sensitisation (from scheduled or unscheduled SPT) by 18 months of age for the protocolised panel of allergens. Each cell gives positive/tested (%).

| **Allergen** | **aP**<br>N = 486 | **wP**<br>N = 486 | **Overall**<br>N = 972 |
|---|---|---|---|
| Wheal size >1mm vs. neg control |  |  |  |
| *D.pteronyssinus* | 14/470 (3.0%) | 15/470 (3.2%) | 29/940 (3.1%) |
| Cat dander | 9/470 (1.9%) | 10/471 (2.1%) | 19/941 (2.0%) |
| Perennial ryegrass | 6/470 (1.3%) | 6/471 (1.3%) | 12/941 (1.3%) |
| Egg | 44/471 (9.3%) | 50/471 (11%) | 94/942 (10%) |
| Cashew | 15/470 (3.2%) | 12/470 (2.6%) | 27/940 (2.9%) |
| Cow's milk | 18/469 (3.8%) | 17/471 (3.6%) | 35/940 (3.7%) |
| Peanut | 20/470 (4.3%) | 21/469 (4.5%) | 41/939 (4.4%) |
| Wheal size ≥3mm vs neg. control |  |  |  |
| *D.pteronyssinus* | 10/470 (2.1%) | 14/470 (3.0%) | 24/940 (2.6%) |
| Cat dander | 2/470 (0.4%) | 5/471 (1.1%) | 7/941 (0.7%) |
| Perennial ryegrass | 3/470 (0.6%) | 2/471 (0.4%) | 5/941 (0.5%) |
| Egg | 33/471 (7.0%) | 35/471 (7.4%) | 68/942 (7.2%) |
| Cashew | 12/470 (2.6%) | 9/470 (1.9%) | 21/940 (2.2%) |
| Cow's milk | 15/469 (3.2%) | 14/471 (3.0%) | 29/940 (3.1%) |
| Peanut | 12/470 (2.6%) | 14/469 (3.0%) | 26/939 (2.8%) |

### Plasma studies

Total IgE and allergen-specific IgE concentrations were similar between groups at 19 months old (Table 5 and Supplementary material 2, Figure E9). Estimates of GMR and the proportions exceeding threshold concentrations were generally imprecise, with little evidence of a difference between the groups.

**Table 5:** Estimated geometric mean ratio (GMR) of total or allergen-specific IgE (kU/L) and percentage point difference in quantitation (IgE ≥ 0.01 kU/L) and sensitisation (IgE ≥ 0.1 kU/L) at 19-months of age, wP versus aP.

|  | **aP**<br>N = 75 |  |  |  | **wP**<br>N = 75 |  |  |  | Median (95% CrI) |  |  |
|---|---|---|---|---|---|---|---|---|---|---|---|
| IgE type | N | GMC | ≥0.01 | ≥0.1 | N | GMC | ≥0.01 | ≥0.1 | GMR | Diff. ≥0.01 | Diff. ≥0.1 |
| Total IgE | 62 | 17.64 | - | - | 53 | 20.43 | - | - | 1.15 (0.70, 1.95) | - | - |
| Tetanus-toxoid | 65 | 0.02 | 65% | 26% | 55 | 0.02 | 64% | 31% | 1.04 (0.42, 2.54) | -1 (-18, 16) | 4 (-11, 21) |
| Egg white | 64 | 0.03 | 75% | 22% | 54 | 0.04 | 74% | 28% | 1.16 (0.57, 2.31) | -1 (-17, 14) | 6 (-9, 21) |
| Whole egg | 65 | 0.03 | 58% | 26% | 55 | 0.03 | 71% | 27% | 1.22 (0.53, 2.74) | 12 (-5, 29) | 1 (-15, 17) |
| Peanut | 64 | 0.03 | 89% | 12% | 55 | 0.04 | 93% | 16% | 1.12 (0.64, 1.90) | 3 (-7, 13) | 3 (-8, 17) |
| Cashew | 64 | 0.01 | 45% | 6% | 53 | 0.01 | 45% | 9% | 0.90 (0.50, 1.61) | 0 (-17, 17) | 3 (-6, 13) |
| Cow’s milk | 62 | 0.05 | 98% | 21% | 53 | 0.06 | 100% | 21% | 1.08 (0.61, 1.95) | 1 (-3, 5) | 0 (-14, 14) |
| D. pteronyssinus | 60 | 0.02 | 97% | 3% | 47 | 0.02 | 98% | 2% | 1.00 (0.77, 1.27) | 1 (-5, 7) | -1 (-7, 5) |
| Cat | 60 | 0.01 | 37% | 2% | 47 | 0.01 | 34% | 2% | 0.82 (0.50, 1.31) | -3 (-19, 15) | 0 (-5, 6) |
| Rye grass | 58 | 0.01 | 52% | 2% | 47 | 0.01 | 45% | 4% | 0.93 (0.55, 1.56) | -6 (-25, 12) | 2 (-4, 9) |

### Safety

There were 154 AEs documented (135 unsolicited, one AESI, and 18 SAEs); most were identified among the subset monitored closely for events (Table 6). Infants in the mixed wP/aP group experienced more AEs compared with those in the aP-only group. Three non-serious AEs in the aP-only group (two with injection site haematoma; one with rash) and four in the mixed wP/aP group (three with injection site haematoma; one with rash) were deemed related to the study vaccines/ trial procedures.

**Table 6:** Number and proportion of infants with any adverse events and the number of adverse events per infant for all participants stratified by receipt of intensive monitoring.

| Characteristic | First 300 PCH |  |  | Remaining |  |  |
|---|---|---|---|---|---|---|
|  | **aP**<br>N = 150 | **wP**<br>N = 150 | **Overall**<br>N = 300 | **aP**<br>N = 336 | **wP**<br>N = 336 | **Overall**<br>N = 672 |
| *Adverse events* |  |  |  |  |  |  |
| Any AE | 41 (27%) | 66 (44%) | 107 (36%) | 4 (1%) | 6 (2%) | 10 (1%) |
| Any related AE | 2 (1%) | 4 (3%) | 6 (2%) | 0 (0%) | 1 (0%) | 1 (0%) |
| Any severe AE | 4 (3%) | 2 (1%) | 6 (2%) | 1 (0%) | 2 (1%) | 3 (0%) |
| Number of AEs |  |  |  |  |  |  |
| 0 | 109 (73%) | 84 (56%) | 193 (64%) | 332 (99%) | 330 (98%) | 662 (99%) |
| 1 | 32 (21%) | 49 (33%) | 81 (27%) | 4 (1%) | 5 (1%) | 9 (1%) |
| 2 | 7 (5%) | 14 (9%) | 21 (7%) | 0 (0%) | 1 (0%) | 1 (0%) |
| 3 | 2 (1%) | 1 (1%) | 3 (1%) | 0 (0%) | 0 (0%) | 0 (0%) |
| 4 | 0 (0%) | 1 (1%) | 1 (0%) | 0 (0%) | 0 (0%) | 0 (0%) |
| 5+ | 0 (0%) | 1 (1%) | 1 (0%) | 0 (0%) | 0 (0%) | 0 (0%) |
| Number of related AEs |  |  |  |  |  |  |
| 0 | 148 (99%) | 146 (97%) | 294 (98%) | 336 (100%) | 335 (100%) | 671 (100%) |
| 1 | 1 (1%) | 4 (3%) | 5 (2%) | 0 (0%) | 1 (0%) | 1 (0%) |
| 2 | 1 (1%) | 0 (0%) | 1 (0%) | 0 (0%) | 0 (0%) | 0 (0%) |
| *Serious adverse events* |  |  |  |  |  |  |
| Any SAE | 4 (3%) | 4 (3%) | 8 (3%) | 2 (1%) | 5 (1%) | 7 (1%) |
| Any related SAE | 0 (0%) | 0 (0%) | 0 (0%) | 0 (0%) | 1 (0%) | 1 (0%) |
| Number of SAEs |  |  |  |  |  |  |
| 0 | 146 (97%) | 146 (97%) | 292 (97%) | 334 (99%) | 331 (99%) | 665 (99%) |
| 1 | 4 (3%) | 3 (2%) | 7 (2%) | 2 (1%) | 4 (1%) | 6 (1%) |
| 2 | 0 (0%) | 0 (0%) | 0 (0%) | 0 (0%) | 1 (0%) | 1 (0%) |
| 3 | 0 (0%) | 1 (1%) | 1 (0%) | 0 (0%) | 0 (0%) | 0 (0%) |

A total of 18 SAEs were reported during the study period (six among six participants in the aP-only group, and 12 SAEs among eight participants in the mixed wP/aP group). A hospital admission for moderate irritability after the 6-week vaccine doses (length of stay < 48 hours) was assessed to be likely related to the wP vaccine; the 17 remaining SAEs were determined to be unrelated to the study vaccines (Supplementary material 2, Table E7). One AESI, laboratory confirmed pertussis infection, was reported in the aP-only group 479 days post-randomisation.

## Discussion

Substituting the first aP vaccine dose at age six weeks old with a wP-containing vaccine is unlikely to result in a reduction in the risk of IgE-mediated food allergy large enough to influence policy. Owing to the greater reactogenicity of wP compared to aP vaccines, we pre-specified a target effect of at least a 3-percentage point reduction in food allergy, or one case of food allergy prevented for every 34 infants vaccinated. The 95% CrI of −0.02 to 0.04 indicates that an effect of this magnitude was unlikely, although a smaller benefit (or harm) cannot be excluded. The prevalence of eczema and skin prick sensitisation by age 12 months was also similar between the aP-only and mixed wP/aP groups, providing further evidence that wP is unlikely to have a material impact on the early onset of atopic diseases in our setting. Importantly, we were not powered to compare schedules for any difference in pertussis protection which is the basis for WHO’s preferential recommendation for wP in countries still using wP.[17]

Our findings are similar to previous trials evaluating the immunomodulatory effects of BCG vaccination at birth which also failed to demonstrate any protection against food allergy, atopic sensitisation, or eczema by age 12 months, although a reduction in the risk of persistent eczema later in childhood has been suggested.[43–45]

In our previous studies we reported that among Australian children born in the late 1990s, those whose first pertussis vaccine was wP had a reduced risk of food allergy (OR; 0.77; 95% CI, 0.62–0.95) and admissions for food anaphylaxis (hazard ratio; 95% 0.47; 95% CI, 0.26–0.83) than those who received aP.[25,46] We undertook this blinded randomised controlled trial because those studies were observational, so we could not exclude the possibility that the observed effects were confounded by differences in baseline risk of food allergy or in health seeking behaviour among infants who received wP rather than aP.

Although the difference in results may be explained by residual bias in the earlier studies, it is important to note factors that limit transportability of the current trial results to the historical cohort examined in those studies. First, there have been important changes in infant feeding practices; in the present trial, most parents reported the introduction of egg, cow’s milk, and peanut before 12 months old in line with current recommendations. While we had no data on feeding practice in our previous studies, Australian and international guidelines at the time recommended delayed introduction of potentially allergenic foods.[47] Earlier introduction of allergenic foods may explain why the incidence of food allergy in the aP-only group (5.3%) was lower than anticipated (10%), albeit broadly consistent with findings from more recent Australian cohorts. For example, the incidence of egg allergy (4.0%) fell between the estimates from other contemporaneous cohorts of Australian children (2.8% and 7.6%),[5,43,48] and the prevalence of peanut allergy (2.0%) was similar to that reported in another randomised trial (2.6%) and a recent observational study (2.6%).[6,43]

Secondly, variability in wP vaccine manufacturing and composition are known to influence vaccine immunogenicity and reactogenicity,[49,50] so it is plausible that any effects on immune phenotype may also be vaccine specific. Our findings cannot necessarily be generalised to other wP vaccine formulations, including the one used previously in Australia.

We found no clinically relevant differences in TT-IgE or egg white-specific IgE responses between treatment groups at 7 or 19 months of age.[46] The absence of effect on IgE responses also differs from previous work from our group showing that wP-priming attenuates Th_2_-dependent bystander immune stimulation in toddlers.[24] Unlike earlier studies, most infants in the current trial were exposed to maternal pertussis vaccination in pregnancy which might influence infant immune responses to wP vaccine in early infancy through Th_2_ skewing or immune interference mediated by high concentrations of transplacentally acquired maternal antibodies.[51,52] Any impact of these mechanisms could not be determined in the current cohort because pre-vaccination blood samples were not collected. It is also possible that the timing of administration may have missed a critical window for immune training or epigenetic modifications.

Our study has several strengths. First, we conducted a closely monitored randomised comparison with blinded assessment of the primary outcome by clinical allergists, using open OFCs and other standardised diagnostic measures. Second, the study included a geographically diverse population of Australian-born infants, with high retention rates and minimal missing data on the primary outcome. Third, 300 infants underwent close safety monitoring and oversight, contributing robust evidence of the tolerability of 5-in-1 wP combination vaccine in infants aged 6 weeks old.

However, our study had limitations that temper our conclusions. First, OFCs were not conducted in all infants with highly probable food allergy and transient cases of IgE-mediated food allergy and eczema occurring before 12 months may have been missed, although unscheduled visits were permitted to maximise ascertainment of these events. While it is likely there was some underascertainment of the primary outcome, there is no reason to expect any misclassification was differential between groups. Second, blood sampling timepoints were designed to support assessment of atopic outcomes without overburdening participants. Therefore, pre-vaccination samples were not collected to assess the contribution of baseline cytokine profiles and transplacentally derived antibodies to subsequent infant immune responses. Finally, we were forced to terminate enrolment early due to inability to procure further doses of the wP combination vaccine. At the last analysis the predicted probability of eventual trial success was very low (<0.01) meaning the trial would likely have stopped early for futility even if further doses could have been procured. As a result, it is unlikely that early stopping would have affected our conclusions.

Despite these limitations, given the lack of evidence of a benefit for any endpoint or in any subgroup, we conclude that a novel mixed wP/aP schedule would be unlikely to achieve a policy-relevant reduction in the prevalence of IgE-mediated food allergy.

## Supporting information

CONSORT checklist

## Data Availability

Data from the study will be available at the completion of follow-up, analysis and reporting of OPTIMUM stage two; no end date. Deidentified, individual participant data and a data dictionary defining each field in the set, will be made available to researchers who provide a methodologically sound proposal to the University of Sydney, Australia (Ms Nicole Wong, Clinical Trial Support Lead; Department: Pro Vice-Chancellor, Research;), subject to a signed data access agreement and any necessary ethics approvals (see ANZCTR trial registration). The study protocol and statistical analysis plan are published. Other study related documentation is available on request.

## Acknowledgement

We acknowledge Aboriginal and Torres Strait Islander peoples as the Traditional Custodians of the lands on which the OPTIMUM study was conducted. We seek their wisdom in our work to improve the health and development of all children.

We thank the 972 families who took part in OPTIMUM and the research nurses, allergists and coordinators at the four participating sites, whose work is reflected in the 96% completion of scheduled follow-up. We would also like to express our gratitude to Kylie Rogers (Data manager, The Kids Research Institute Australia), Dr Andrew McLean-Tooke (Head, PathWest Immunology QEII campus, Western Australia, Australia) for their valuable contributions, and to the independent DSMC for their detailed oversight.

The authors wish to dedicate this work to the memory of Professor Katrina (Katie) Allen AO, who passed away peacefully in January 2026. We express our gratitude for her outstanding contributions to the field of paediatric allergy and her invaluable contribution to the development of the OPTIMUM study National Health and Medical Research Council grant proposal.

## Abbreviations

AE: Adverse event
AESI: Adverse event of special interest
aP: Acellular pertussis vaccine
ASCIA: Australasian Society of Clinical Immunology and Allergy
CI: Confidence interval
CrI: Credible interval
DSMC: Data Safety and Monitoring Committee
DTaP-HepB-IPV-Hib: Diphtheria, tetanus, aP, hepatitis B, inactivated poliovirus types 1, 2, and 3, and *Haemophilus influenzae* type b
DTwP-Hib-HepB: Diphtheria, tetanus, whole-cell pertussis, *Haemophilus influenzae* type b, hepatitis B vaccine
IPV: Inactivated poliovirus vaccine
NIP: National Immunisation Program
SAE: Serious adverse event
SPT: Skin prick test
TT: Tetanus toxoid
OFC: Oral food challenge
OR: Odds ratio
WHO: World Health Organization
wP: Whole-cell pertussis vaccine

## References

1. Osborne NJ, Koplin JJ, Martin PE, Gurrin LC, Lowe AJ, Matheson MC, et al. Prevalence of challenge-proven IgE-mediated food allergy using population-based sampling and predetermined challenge criteria in infants. J Allergy Clin Immunol. 2011;127: 668–676.e2. doi:10.1016/j.jaci.2011.01.039

2. Mullins RJ, Dear KBG, Tang MLK. Changes in Australian food anaphylaxis admission rates following introduction of updated allergy prevention guidelines. J Allergy Clin Immunol. 2022;150: 140–145.e1. doi:10.1016/j.jaci.2021.12.795

3. Joshi PA, Smith J, Vale S, Campbell DE. The Australasian Society of Clinical Immunology and Allergy infant feeding for allergy prevention guidelines. Med J Aust. 2019;210: 89–93. doi:10.5694/mja2.12102

4. Ierodiakonou D, Garcia-Larsen V, Logan A, Groome A, Cunha S, Chivinge J, et al. Timing of allergenic food introduction to the infant diet and risk of allergic or autoimmune disease: A systematic review and meta-analysis. JAMA. 2016;316: 1181–1192. doi:10.1001/jama.2016.12623

5. Koplin JJ, Shifti DM, Soriano VX, Wijesuriya R, Moreno-Betancur M, Odoi A, et al. Egg allergy prevalence before and after guidelines for earlier egg introduction. JAMA Pediatrics. 2026. doi:10.1001/jamapediatrics.2026.2080

6. Soriano VX, Peters RL, Moreno-Betancur M, Ponsonby A-L, Gell G, Odoi A, et al. Association between earlier introduction of peanut and prevalence of peanut allergy in infants in Australia. JAMA. 2022;328: 48–56. doi:10.1001/jama.2022.9224

7. Yu W, Freeland DMH, Nadeau KC. Food allergy: Immune mechanisms, diagnosis and immunotherapy. Nat Rev Immunol. 2016;16: 751–765. doi:10.1038/nri.2016.111

8. Pérez Chacón G, Ramsay J, Brennan-Jones CG, Estcourt MJ, Richmond P, Holt P, et al. Whole-cell pertussis vaccine in early infancy for the prevention of allergy in children. Cochrane Database Syst Rev. 2021;9: CD013682. doi:10.1002/14651858.CD013682.pub2

9. Keja K, Chan C, Hayden G, Henderson RH. Expanded programme on immunization. World Health Stat Q. 1988;41: 59–63.

10. Murphy S, Kelly A, Kelly DF. Global variations in immunisation strategies against pertussis in infancy. Expert Rev Vaccines. 2025;24: 882–903. doi:10.1080/14760584.2025.2562197

11. DTP-containing vaccine boosters programme information. Available: https://www.gavi.org/our-support/guidelines/dtp-containing-vaccine-boosters-programme-information

12. Xing D, Corbel M, Dobbelaer R, Knezevic I. WHO working group on standardisation and control of acellular pertussis vaccines—report of a meeting held on 16–17 March 2006, St. Albans, United Kingdom. Vaccine. 2007;25: 2749–2757. doi:10.1016/j.vaccine.2006.09.050

13. Decker MD, Edwards KM. Pertussis (whooping cough). J Infect Dis. 2021;224: S310–S320. doi:10.1093/infdis/jiaa469

14. McIntyre P, Forrest J, Heath T, Burgess M, Harvey B. Pertussis vaccines: Past, present and future in Australia. 1998;22: 125–32.

15. Torvaldsen S, Simpson JM, McIntyre PB. Effectiveness of pertussis vaccination in New South Wales, Australia, 1996–1998. Eur J Epidemiol. 2003;18: 63–69. doi:10.1023/a:1022588118030

16. Diavatopoulos DA, Mills KH, Kester KE, Kampmann B, Silerova M, Heininger U, et al. PERISCOPE: Road towards effective control of pertussis. Lancet Infect Dis. 2019;19: e179–e186. doi:10.1016/S1473-3099(18)30646-7

17. World Health Organization. Pertussis vaccines: WHO position paper, august 2015—recommendations. Vaccine. 2016;34: 1423–1425. doi:10.1016/j.vaccine.2015.10.136

18. Silva Antunes R da, Babor M, Carpenter C, Khalil N, Cortese M, Mentzer AJ, et al. Th1/Th17 polarization persists following whole-cell pertussis vaccination despite repeated acellular boosters. J Clin Invest. 2018;128: 3853–3865. doi:10.1172/JCI121309

19. Iwasaki A, Omer SB. Why and how vaccines work. Cell. 2020;183: 290–295. doi:10.1016/j.cell.2020.09.040

20. Aalberse RC, Gruber C, Ljungman M, Kakat S, Wahn U, Niggemann B, et al. Further investigations of the IgE response to tetanus and diphtheria following covaccination with acellular rather than cellular *Bordetella pertussis*. Pediatr Allergy Immunol. 2019;30: 841–847. doi:10.1111/pai.13113

21. Nilsson L, Gruber C, Granstrom M, Bjorksten B, Kjellman NI. Pertussis IgE and atopic disease. Allergy. 1998;53: 1195–1201.

22. Silva Antunes R da, Soldevila F, Pomaznoy M, Babor M, Bennett J, Tian Y, et al. A system-view of *Bordetella pertussis* booster vaccine responses in adults primed with whole-cell versus acellular vaccine in infancy. JCI Insight. 2021;6. doi:10.1172/jci.insight.141023

23. Pérez Chacón G, Estcourt MJ, Totterdell J, Marsh JA, Perrett KP, Campbell DE, et al. Immunogenicity, reactogenicity, and IgE-mediated immune responses of a mixed whole-cell and acellular pertussis vaccine schedule in Australian infants: A randomised, double-blind, noninferiority trial. PLoS Med. 2024;21: e1004414. doi:10.1371/journal.pmed.1004414

24. Holt PG, Snelling T, White OJ, Sly PD, DeKlerk N, Carapetis J, et al. Transiently increased IgE responses in infants and pre-schoolers receiving only acellular diphtheria-pertussis-tetanus (DTaP) vaccines compared to those initially receiving at least one dose of cellular vaccine (DTwP) - immunological curiosity or canary in the mine? Vaccine. 2016;34: 4257–4262. doi:10.1016/j.vaccine.2016.05.048

25. Estcourt MJ, Campbell DE, Gold MS, Richmond P, Allen KJ, Quinn HE, et al. Whole-cell pertussis vaccination and decreased risk of IgE-mediated food allergy: A nested case-control study. J Allergy Clin Immunol Pract. 2020;8: 2004–2014. doi:10.1016/j.jaip.2019.12.020

26. Pérez Chacón G, Estcourt MJ, Totterdell J, Campbell DE, Perrett KP, Marsh JA, et al. OPTIMUM study protocol: An adaptive randomised controlled trial of a mixed whole-cell/acellular pertussis vaccine schedule. BMJ Open. 2020;10: e042838. doi:10.1136/bmjopen-2020-042838

27. Totterdell JA, Pérez Chacón G, Estcourt MJ, Jones M, Richmond P, Snelling TL, et al. Statistical analysis plan for the OPTIMUM study: Optimising immunisation using mixed schedules, an adaptive randomised controlled trial of a mixed whole-cell/acellular pertussis vaccine schedule. Trials. 2022;23: 121. doi:10.1186/s13063-021-05874-6

28. Rusmil K, Gunardi H, Fadlyana E, Dhamayanti M, Sekartini R, Satari HI, et al. The immunogenicity, safety, and consistency of an indonesia combined DTP-HB-Hib vaccine in expanded program on immunization schedule. BMC Pediatrics. 2015;15: 219. doi:10.1186/s12887-015-0525-2

29. Eradicate polio now | global polio eradication | GPEI home. Available: https://polioeradication.org/

30. Pérez Chacón G, McAlister S, Totterdell J, Estcourt MJ, Marsh JA, Jones M, et al. Immune impacts of infant whole-cell and acellular pertussis vaccination on co-administered vaccines. J Infect. 2025;91: 106515. doi:10.1016/j.jinf.2025.106515

31. Severity scoring of atopic dermatitis: the SCORAD index. Consensus Report of the European Task Force on Atopic Dermatitis. Dermatology. 1993;186: 23–31. doi:10.1159/000247298

32. Australasian Society of Clinical Immunology and Allergy. Cow’s milk challenge protocol. 2014.

33. Australasian Society of Clinical Immunology and Allergy. Nut challenge protocol (single nut). 2014.

34. Australasian Society of Clinical Immunology and Allergy. Nut challenge protocols - Appendix. 2016.

35. Australasian Society of Clinical Immunology and Allergy. Peanut butter challenge protocol. 2014.

36. Australasian Society of Clinical Immunology and Allergy. Cooked egg challenge protocol. 2014.

37. Sampson HA, Gerth van Wijk R, Bindslev-Jensen C, Sicherer S, Teuber SS, Burks AW, et al. Standardizing double-blind, placebo-controlled oral food challenges: American Academy of Allergy, Asthma & Immunology-European Academy of Allergy and Clinical Immunology PRACTALL consensus report. J Allergy Clin Immunol. 2012;130: 1260–1274. doi:10.1016/j.jaci.2012.10.017

38. National Health and Medical Research Council. Guidance: Safety monitoring and reporting in clinical trials involving therapeutic goods. Canberra: National Health; Medical Research Council; 2016.

39. Brown EG, Wood L, Wood S. The medical dictionary for regulatory activities (MedDRA). Drug Saf. 1999;20: 109–117. doi:10.2165/00002018-199920020-00002

40. R Core Team. R: A language and environment for statistical computing. Vienna, Austria: R Foundation for Statistical Computing; 2024. Available: https://www.R-project.org/

41. Gabry J, Češnovar R, Johnson A, Bronder S. Cmdstanr: R interface to ‘CmdStan’. 2026. Available: https://github.com/stan-dev/cmdstanr

42. Stan reference manual. 2026. Available: https://mc-stan.org

43. Messina NL, Gardiner K, Pittet LF, Forbes EK, Francis KL, Freyne B, et al. Neonatal BCG vaccination for prevention of allergy in infants: The MIS BAIR randomised controlled trial. Clin Exp Allergy. 2024;54: 682–693. doi:10.1111/cea.14537

44. Pittet LF, Messina NL, Gardiner K, Freyne B, Abruzzo V, Francis KL, et al. Prevention of infant eczema by neonatal Bacillus Calmette-Guérin vaccination: The MIS BAIR randomized controlled trial. Allergy. 2022;77: 956–965. doi:10.1111/all.15022

45. Pittet LF, Messina NL, Francis KL, Donath S, Flanagan KL, Forbes EK, et al. Neonatal Bacillus Calmette-Guérin Vaccination Decreases Eczema Incidence at 5 years: The MIS BAIR Randomised Controlled Trial. Allergy. 2025;80: 3444–3446. doi:10.1111/all.16677

46. Pérez Chacón G, Fathima P, Jones M, Estcourt MJ, Gidding HF, Moore HC, et al. Infant whole-cell versus acellular pertussis vaccination in 1997 to 1999 and risk of childhood hospitalization for food-induced anaphylaxis: Linked administrative databases cohort study. J Allergy Clin Immunol Pract. 2024;12: 670–680. doi:10.1016/j.jaip.2023.12.048

47. Fiocchi MA, Assa’ad MA, Bahna MS, et al. Food allergy and the introduction of solid foods to infants: A consensus document. Ann Allergy Asthma Immunol. 2006;97: 10–21. doi:10.1016/s1081-1206(10)61364-6

48. Walker SVM, D’Vaz N, Pretorius RA, Lo J, Christophersen C, Prescott SL, et al. Infant diet recommendations reduce IgE-mediated egg, peanut, and cow’s milk allergies. J Allergy Clin Immunol Pract. 2025;13: 3077–3083.e1. doi:10.1016/j.jaip.2025.06.012

49. Zhang L, Prietsch SOM, Axelsson I, Halperin SA. Acellular vaccines for preventing whooping cough in children. Cochrane Database Syst Rev. 2014;2014: CD001478. doi:10.1002/14651858.CD001478.pub6

50. Alghounaim M, Alsaffar Z, Alfraij A, Bin-Hasan S, Hussain E. Whole-cell and acellular pertussis vaccine: Reflections on efficacy. Med Princ Pract. 2022;31: 313–321. doi:10.1159/000525468

51. Sereeyothin C, Kerr SJ, Poovorawan Y, Maertens K, Wanlapakorn N. The effect of preexisting antibodies from Tdap vaccination during pregnancy on infant antibody responses to the pertussis-containing vaccines. Hum Vaccin Immunother. 2025;21: 2575638. doi:10.1080/21645515.2025.2575638

52. Saso A, Kanteh E, Jeffries D, Okoye M, Mohammed N, Kumado M, et al. The effect of pertussis vaccination in pregnancy on the immunogenicity of acellular or whole-cell pertussis vaccination in Gambian infants (GaPS): a single-centre, randomised, controlled, double-blind, phase 4 trial. Lancet Infect Dis. 2025;25: 909–924. doi:10.1016/S1473-3099(25)00072-6

