## Supplementary material 1 for "Efficacy of an early first dose of whole-cell pertussis vaccine in infants for the prevention of IgE-mediated food allergy: A randomised controlled trial"

2026-11-06

Checklist

- ☐ Vital Signs
- ☐ Physical Examination
- ☐ Food Challenge
- ☐ Concomitant Medications
- ☐ SCORAD

*Study Team Members:*

*Data Entry Complete* ☐

*Allergy Report required?* ☐ *Yes* ☐ *No*

Study Staff Initials:_______

Date: _____ /_______ /_______

#### Physical Examination

Weight (kg): ________ Height (cm): ________

| BODY SYSTEM | Normal | Abnormal | CS? | Not Done | If Abnormal/Not Done, please comment |
|---|---|---|---|---|---|
| General appearance | ☐ | ☐ | ☐ Yes ☐ No | ☐ | |
| Head and Neck | ☐ | ☐ | ☐ Yes ☐ No | ☐ | |
| Ears, Nose and Throat | ☐ | ☐ | ☐ Yes ☐ No | ☐ | |
| Eyes | ☐ | ☐ | ☐ Yes ☐ No | ☐ | |
| Complete skin examination | ☐ | ☐ | ☐ Yes ☐ No | ☐ | |
| Cardiovascular system | ☐ | ☐ | ☐ Yes ☐ No | ☐ | |
| Respiratory system | ☐ | ☐ | ☐ Yes ☐ No | ☐ | |
| Abdominal system | ☐ | ☐ | ☐ Yes ☐ No | ☐ | |
| Nervous system | ☐ | ☐ | ☐ Yes ☐ No | ☐ | |
| Other: __________________ | ☐ | ☐ | ☐ Yes ☐ No | ☐ | |
| Additional Comments: | | | | | |

Baseline vitals to be done before the food challenge. Can be repeated anytime during food challenge, as judged necessary by the Investigator or supervising study doctor

*Measurement after the subject has been sitting for approximately 5 minutes.*

**BASELINE ASSESSMENT**

Weight: [ ][ ].[ ][ ] Kg

Height: [ ][ ][ ].[ ] cm

Temperature: [ ][ ] . [ ] °C

Pulse Rate: [ ][ ][ ] beat/min

Blood Pressure: [ ][ ][ ] / [ ][ ] mmHg

Respiratory Rate: [ ][ ] breaths/min

$SaO_2$: [ ][ ][ ] %

**Record any pre-existing rash**

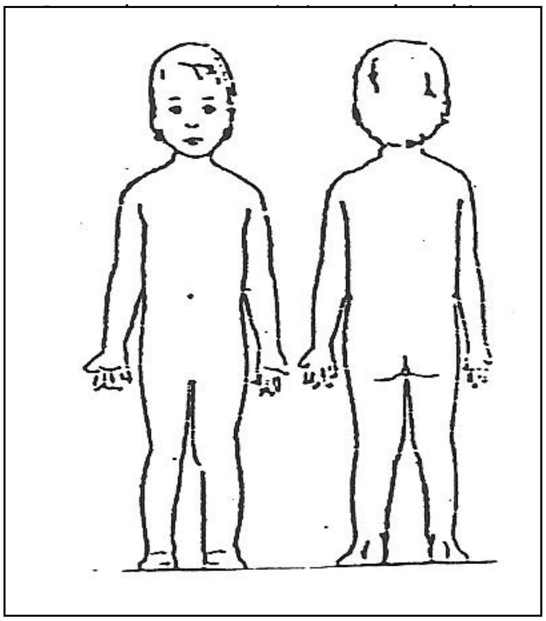

Air Entry: ______________________________________________

Skin: ______________________________________________

A – EXTENT - Percentage of Body Surface Area involved (%): _________

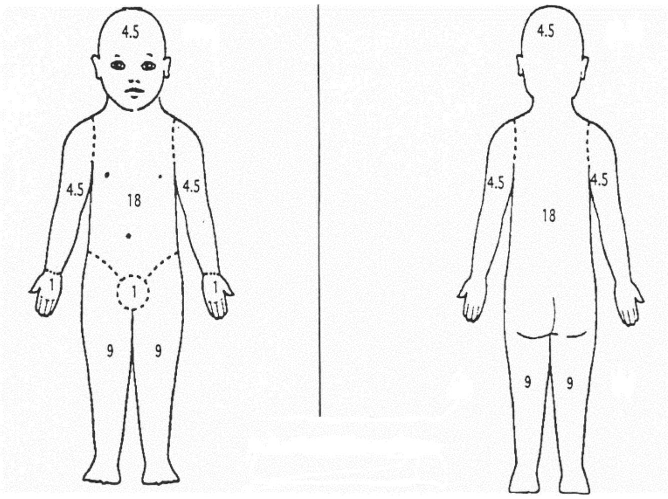

B – INTENSITY

| | SCORE | | | |
|---|---|---|---|---|
| | **0** | **1** | **2** | **3** |
| Intensity of erythema / darkening | ☐ Absence | ☐ Mild | ☐ Mod | ☐ Severe |
| Intensity of edema / papulation/ swelling | ☐ Absence | ☐ Mild | ☐ Mod | ☐ Severe |
| Intensity of oozing / crust | ☐ Absence | ☐ Mild | ☐ Mod | ☐ Severe |
| Intensity of excoriation / scratch marks | ☐ Absence | ☐ Mild | ☐ Mod | ☐ Severe |
| Intensity of lichenification / prurigo / skin thickening | ☐ Absence | ☐ Mild | ☐ Mod | ☐ Severe |
| Intensity of dryness (dryness is evaluated on uninvolved areas) | ☐ Absence | ☐ Mild | ☐ Mod | ☐ Severe |

Other:
_______________________________________________
_______________________________________________
_______________________________________________
_______________________________________________

#### ***Allergen***

| | | | | | | | | | |
|---|---|---|---|---|---|---|---|---|---|
| Allergen being challenged: | | | ☐ Peanut<br>☐Cashew<br>☐Egg<br>☐Milk<br>☐Other | | | | | | |
| Last skin prick test (SPT) to allergen (mm):<br>(From OPTIMUM or most recent SPT) |  |  |  |  | Date SPT: | --------/---------/-- |  |  |  |
| Previous reaction to allergen: | Yes | ☐ | No (avoidance based on SPT) | | | ☐ | | | |
| Worst documented reaction to allergen | | | | | | | | | |
| IgE Mediated Anaphylaxis | ☐ | | IgE-mediated generalised allergic reaction | | ☐ | | | | |
| Never had | ☐ | | Eczema flare | | ☐ | | | | |
| Other: | | | | | | | | | |
| Last clinical reaction to allergen: | | | | | | | | | |
| < 6 months | ☐ | 6-12 mo ago | ☐ | 12-24 mo ago | ☐ | > 24 months | ☐ | Never had | ☐ |
| Has child had allergen in question since booking: ☐Yes ☐No | | | | | | | | | |
| If yes, did the child react: ☐Yes * ☐No<br>**If yes, please record reaction on Food Allergy History form. Unless counter to clinician recommendation, challenge to proceed irrespective of answer.* | | | | | | | | | |

### Oral food challenge SOP

- Please record in the table below details for each dose of the food given
- **20 minute intervals** between doses. Can be extended upon clinician discretion
- Dose can be repeated upon clinical discretion
- Challenge food can be mixed with other non-allergenic food upon clinical discretion
- Objective symptoms that stop a challenge should be treated as a safety precaution

DELETE FOOD AS APPROPRIATE

**PEANUT PASTE ORAL FOOD CHALLENGE**

**Bega smooth peanut paste to be used.**

| Cumulative dose (if all consumed) | Measured Dose | Dose consumed | Start time dose given (24 hour clock) |
|---|---|---|---|
| Trace | touch inside lip (not to touch outside lip) | ☐ All | (HH:MM): __________ |
| 1/16 tsp | ☐ 1/16 tsp | ☐ All<br>☐ _________<br>(best estimate in tsp) | (HH:MM): __________ |
| 3/16 tsp | ☐ 1/8 tsp | ☐ All<br>☐ _________<br>(best estimate in tsp) | (HH:MM): __________ |
| 7/16 tsp | ☐ ¼ tsp | ☐ All<br>☐ _________<br>(best estimate in tsp) | (HH:MM): __________ |
| 15/16 tsp | ☐ ½ tsp | ☐ All<br>☐ _________<br>(best estimate in tsp) | (HH:MM): __________ |
| 1 15/16 tsp | ☐ 1 tsp | ☐ All<br>☐ _________<br>(best estimate in tsp) | (HH:MM): __________ |

Cumulative Dose (*Sum of all doses effectively given*): __________ tsp

Eliciting dose: __________ tsp

(*last dose given before reaction- *Note, if more than ½ of dose given, record as ED. If less than half of dose given, consider ED to be the preceding dose)*

**CASHEW PASTE ORAL FOOD CHALLENGE**

**MACRO Natural Spread brand cashew paste to be used**

| Cumulative dose (if all consumed) | Measured Dose | Dose consumed | Start time dose given (24 hour clock) |
|---|---|---|---|
| Trace | smear inside lip (not to touch outside lip) | ☐ All<br>☐ _________<br>(best estimate in tsp) | (HH:MM): __________ |
| 1/16 tsp | ☐ 1/16 tsp | ☐ All<br>☐ _________<br>(best estimate in tsp) | (HH:MM): __________ |
| 3/16 tsp | ☐ 1/8 tsp | ☐ All<br>☐ _________<br>(best estimate in tsp) | (HH:MM): __________ |
| 7/16 tsp | ☐ ¼ tsp | ☐ All<br>☐ _________<br>(best estimate in tsp) | (HH:MM): __________ |
| 15/16 tsp | ☐ ½ tsp | ☐ All<br>☐ _________<br>(best estimate in tsp) | (HH:MM): __________ |
| 1 15/16 tsp | ☐ 1 tsp | ☐ All<br>☐ _________<br>(best estimate in tsp) | (HH:MM): __________ |

Cumulative Dose (*Sum of all doses effectively given*): __________ tsp
Eliciting dose: __________ tsp
(*last dose given before reaction- *Note, if more than ½ of dose given, record as ED. If less than half of dose given, consider ED to be the preceding dose)*

**SCRAMBLED EGG ORAL FOOD CHALLENGE**

**Preparation instructions**

1 Beat one large egg (yolk and white) with a fork for 1 minute
2 Place in microwave
3 Cook at 10 second intervals, mashing the egg mixture with a fork each time.
4 Cook egg until it is rendered semi-solid (approximately a total of 30 seconds of cooking is sufficient)

| Cumulative dose (if all consumed) | Planned Dose | Dose consumed | Start time dose given (24 hour clock) |
|---|---|---|---|
| Trace | smear inside lip (not to touch outside lip) | ☐ All<br>☐ _________ | (HH:MM): __________ |
| 1/16 tsp | ☐ 1/16 tsp | ☐ All<br>☐ _________<br>(best estimate in tsp) | (HH:MM): __________ |
| 3/16 tsp | ☐ 1/8 tsp | ☐ All<br>☐ _________<br>(best estimate in tsp) | (HH:MM): __________ |
| 7/16 tsp | ☐ ¼ tsp | ☐ All<br>☐ _________<br>(best estimate in tsp) | (HH:MM): __________ |
| 15/16 tsp | ☐ ½ tsp | ☐ All<br>☐ _________<br>(best estimate in tsp) | (HH:MM): __________ |
| 1 15/16 tsp | ☐ 1 tsp | ☐ All<br>☐ _________<br>(best estimate in tsp) | (HH:MM): __________ |
| 2 15/16 tsp | ☐ 2 tsp | ☐ All<br>☐ _________<br>(best estimate in tsp) | (HH:MM): __________ |
| Whole egg | ☐ rest of egg | ☐ All<br>☐ _________<br>(best estimate in tsp) | (HH:MM): __________ |

Cumulative Dose (*Sum of all doses effectively given*): __________ tsp
Eliciting dose: __________ tsp
(*last dose given before reaction- *Note, if more than ½ of dose given, record as ED. If less than half of dose given, consider ED to be the preceding dose)*

**COWS MILK ORAL FOOD CHALLENGE**

CHALLENGE SUBSTANCE

1. Less than 12 months old – cow's milk based infant formula
2. More than 12 months old - full cream cow's milk

| Cumulative dose (if all consumed) | Dose | Dose consumed | Start time dose given (24 hour clock) |
|---|---|---|---|
| 0.1ml | ☐ 0.1 ml | ☐ All<br>☐ _________ (best estimate in mls) | (HH:MM): __________ |
| 1.1 ml | ☐ 1ml | ☐ All<br>☐ _________ (best estimate in mls) | (HH:MM): __________ |
| 6.1 ml | ☐ 5ml | ☐ All<br>☐ _________ (best estimate in mls) | (HH:MM): __________ |
| 21.1ml | ☐ 15ml | ☐ All<br>☐ _________ (best estimate in mls) | (HH:MM): __________ |
| 61.1ml | ☐ 40ml | ☐ All<br>☐ _________ (best estimate in mls) | (HH:MM): __________ |
| 161.1ml | ☐ 100ml | ☐ All<br>☐ _________ (best estimate in mls) | (HH:MM): __________ |

Cumulative Dose (*Sum of all doses effectively given*): __________ mls

Eliciting dose: __________ mls

(*last dose given before reaction- *Note, if more than ½ of dose given, record as ED. If less than half of dose given, consider ED to be the preceding dose)*

Did the subject experience an objective reaction during the food challenge? ☐ Yes ☐ No
If yes, record all details in the table below.

Erythematous rash ☐ Yes ☐ No
If Yes, specify the % Rash area involved: _______ % (Record the Rash Grade below)

| **Objective Symptom** | Time the symptom first appeared (24 hour clock) | Highest severity Grade (refers to ***"Oral Food Challenge Symptom Score Sheet"*** *at the end of this worksheet*) |
|---|---|---|
| Pruritus | (HH:MM): _________ | ☐ 0=Absent ☐ 1=Mild ☐ 2=Moderate ☐ 3=Severe |
| Urticaria | (HH:MM): _________ | ☐ 0=Absent ☐ 1=Mild ☐ **2=Moderate** ☐ **3=Severe** |
| Angioedema | (HH:MM): _________ | ☐ 0=Absent ☐ 1=Mild ☐ **2=Moderate** ☐ **3=Severe** |
| Rash (If Grade ≥ 1, record % above) | (HH:MM): _________ | ☐ 0=Absent ☐ 1=Mild ☐ 2=Moderate ☐ **3=Severe** |
| Sneezing/ Itching | (HH:MM): _________ | ☐ 0=Absent ☐ 1=Mild ☐ 2=Moderate ☐ **3=Severe** |
| Nasal Congestion | (HH:MM): _________ | ☐ 0=Absent ☐ 1=Mild ☐ 2=Moderate ☐ **3=Severe** |
| Rhinorrhea | (HH:MM): _________ | ☐ 0=Absent ☐ 1=Mild ☐ 2=Moderate ☐ **3=Severe** |
| Laryngeal | (HH:MM): _________ | ☐ 0=Absent ☐ 1=Mild ☐ **2=Moderate** ☐ **3=Severe** |
| Wheezing | (HH:MM): _________ | ☐ 0=Absent ☐ **1=Mild** ☐ **2=Moderate** ☐ **3=Severe** |
| Diarrhea | (HH:MM): _________ | ☐ 0=Absent ☐ 1=Mild ☐ **2=Moderate** ☐ **3=Severe** |
| Vomiting | (HH:MM): _________ | ☐ 0=Absent ☐ 1=Mild ☐ **2=Moderate** ☐ **3=Severe** |
| Cardiovascular | (HH:MM): _________ | ☐ 0=Absent ☐ 1=Mild ☐ **2=Moderate** ☐ **3=Severe** |
| Other:_____________ | (HH:MM): _________ | ☐ 0=Absent ☐ 1=Mild ☐ 2=Moderate ☐ 3=Severe |
| Other:_____________ | (HH:MM): _________ | ☐ 0=Absent ☐ 1=Mild ☐ 2=Moderate ☐ 3=Severe |
| Other:_____________ | (HH:MM): _________ | ☐ 0=Absent ☐ 1=Mild ☐ 2=Moderate ☐ 3=Severe |

** Any listed **Bold** reaction: Stop dosing, considered positive food challenge
**3 or more underlined scoring area's: Stop dosing, considered positive food challenge

| | | |
|---|---|---|
| *If cardiovascular grade 1* | Weak | ☐ *Yes* ☐ *No* |
| | Dizzy | ☐ *Yes* ☐ *No* |
| | Tachycardia | ☐ *Yes* ☐ *No* |
| | Color Change | ☐ *Yes* ☐ *No* |
| | Mental status change | ☐ *Yes* ☐ *No* |
| | Mild hypotension (weak rapid pulse and/or 10-20% drop in blood pressure from baseline) | ☐ *Yes* ☐ *No* |
| *If cardiovascular grade 2* | Moderate hypotension (more than 20% drop in BP from baseline) | ☐ *Yes* ☐ *No* |
| | Significant change in mental status | ☐ *Yes* ☐ *No* |
| | Light-headedness | ☐ *Yes* ☐ *No* |
| | Feeling of "pending doom" | ☐ *Yes* ☐ *No* |
| *If cardiovascular grade 3* | Cardiovascular collapse | ☐ *Yes* ☐ *No* |
| | Signs of impaired circulation | ☐ *Yes* ☐ *No* |
| | Unconsciousness | ☐ *Yes* ☐ *No* |
| | Bradycardia | ☐ *Yes* ☐ *No* |
| | Cardiac arrest | ☐ *Yes* ☐ *No* |

Did the subject reach the study specified stopping criteria? ☐ Yes ☐ No

**If no:** Did the subject complete the food challenge? ☐ Yes ☐ No

***Reactions to food challenge <u>should not</u> be recorded as AEs, except in case of SAE.***
***In case of SAE please report "due to food challenge" in the verbatim (i.e Anaphylaxis due to food challenge)***

**Oral Food Challenge Symptom Score Sheet**
**(Displayed by Categories I to V)**

**<u>Stopping criteria</u>**
Any listed **bold** reaction: Stop dosing, considered positive food challenge
3 or more <u>underlined</u> scoring area's: Stop dosing, considered positive food challenge

I. SKIN

A. *Erythematous Rash*- % area involved (see diagram below): ______________________

B. *Pruritus*
- 0 = Absent
- 1 = Mild: occasional scratching
- 2 = Moderate: scratching continuously for >2 minutes at a time
- <u>3 = Severe:</u> hard continuous scratching excoriations

C. *Urticaria/Angioedema*
- 0 = Absent
- <u>1 = Mild:</u> <3 hives, or mild lip edema
- **2 = Moderate**: <10 hives but >3, or significant lip or face edema
  - i. Any hives must persist for > 5 minutes, simultaneously or contemporaneously with other symptoms.
  - ii. The symptoms have to either persist from the previous dose OR have to reoccur.
- **3 = Severe**: generalized involvement+

D. *Rash*
- 0 = Absent
- 1 = Mild: few areas of faint erythema
- <u>2 = Moderate:</u> areas of erythema (>20% and <50%), macular and raised rash
- **3 = Severe:** generalized marked erythema (>50%), extensive raised lesions (>25%)

| | |
|---|---|
| Head | 4.5% |
| Neck | 1% |
| Anterior trunk | 18% |
| Posterior trunk | 18% |
| Leg | 18% |
| Arm | 9% |

*II. UPPER RESPIRATORY*

*A. Sneezing/Itching*
- 0 = Absent
- 1 = Mild: rare bursts
- <u>2 = Moderate</u>*:* bursts <10, intermittent rubbing of nose / eyes / external ear canals
- **3 = Severe**: continuous rubbing of nose / eyes, periocular swelling and/or long bursts of sneezing

*B. Nasal Congestion*
- 0 = Absent
- 1 = Mild: some hindrance to breathing
- <u>2 = Moderate:</u> nostrils feel blocked, breathing through mouth most of time
- **3 = Severe:** nostrils occluded

*C. Rhinorrhea*
- 0 = Absent
- 1 = Mild: occasional sniffling
- <u>2 = Moderate:</u> frequent sniffling, requires tissues
- **3 = Severe:** nose runs freely despite sniffling and tissues

*D. Laryngeal*

0 = Absent

1 = Mild: throat clearing, occasional cough

**2 = Moderate:** hoarseness, frequent dry cough

**3 = Severe:** inspiratory stridor

*III. LOWER RESPIRATORY*

*A. Wheezing*

0 = Absent

**1 = Mild:** expiratory wheezing to auscultation

**2 = Moderate:** dyspnea, inspiratory and expiratory wheezing

**3 = Severe:** dyspnea, use of accessory muscles, audible wheezing

*IV. GASTROINTESTINAL*

*A. Subjective Complaints*

0 = Absent

*1* = Mild: itchy mouth/throat, c/o nausea, abdominal pain, no change in activity

*2* = Moderate: frequent c/o nausea or abdominal pain, decreased activity

*3* = Severe: patient in bed; crying, notably distressed

*B. Objective Complaints*

0 = Absent

*1* = Mild: 1 episode of emesis or diarrhea

**2 = Moderate**: 2-3 episodes of emesis or diarrhea or 1 of each

**3 = Severe:** >3 episodes of emesis or diarrhea or 2 of each

*V. CARDIOVASCULAR/NEUROLOGIC*

0 = Normal: heart rate or BP for age/baseline

*1* = Mild: color change, subjective response (weak, dizzy), or tachycardia, mental status change, mild hypotension (weak rapid pulse and/or 10-20% drop in blood pressure from baseline)

**2 = Moderate:** drop in blood pressure >20% from baseline, significant change in mental status, light-headedness, feeling of “pending doom”

**3 = Severe:** cardiovascular collapse, signs of impaired circulation, unconsciousness, bradycardia, cardiac arrest.

#### Treatments administered during this Food Challenge:

**Weight: kg**

##### Epinephrine

Was Epinephrine administered during this challenge? ☐ Yes ☐ No

If yes,

- How many times has the epinephrine been given? ________
- Enter the Start Time of each epinephrine dose given below:

**Dose of INTRAMUSCULAR adrenaline:**

**0.___ ml (1:1000)**

| | | | | | |
|---|---|---|---|---|---|
| Dose Number: | 1 | Dose Number: | 2 | Dose Number: | 3 |
| Dose Time (HH:MM): | _________ | Dose Time (HH:MM): | _________ | Dose Time (HH:MM): | _________ |
| Dose Amount: | _________ | Dose Amount: | _________ | Dose Amount: | _________ |

Call Rapid Response Team XXXX

##### Antihistamines

Was Antihistamines administered during this challenge? ☐ Yes ☐ No

If yes,

- How many times have Antihistamines been given? ________
- Enter the Start Time of each Antihistamine given below:

**Name of Antihistamine:**
**Dose:**

| | | | | | |
|---|---|---|---|---|---|
| Dose Number: | 1 | Dose Number: | 2 | Dose Number: | 3 |
| Dose Time (HH:MM): | _________ | Dose Time (HH:MM): | _________ | Dose Time (HH:MM): | _________ |
| Dose Amount: | _________ | Dose Amount: | _________ | Dose Amount: | _________ |

##### Steroids

Were Steroids administered during this challenge? ☐ Yes ☐ No

Drug Name: ___________________________ Dose Time (HH:MM): ______________ Dose Amount: _________

##### Other medications

Were other medications administered during this challenge? ☐ Yes ☐ No

Drug Name: ___________________________ Dose Time (HH:MM): ______________ Dose Amount: _________

*All medications administered during this challenge (Epinephrine, Antihistamines, Steroids or Others) must be recorded also in the concomitant medications appropriate log and reported in the Prior and Concomitant Medications page in the eCRF.*

Discharge:

Discharge Date (DD / MMM / YYYY): _____ /_______ /_201___

Discharge Time (HH:MM): __________

***Patient discharged after 2hrs of observation minimum after the last dose ingested***

Observation period extended beyond 2 hours (judged necessary by investigator) ☐ Yes ☐ No

If Adrenaline given hospital policy states patient to remain for observation 4 hours post adrenaline administration

**Reviewed and assessed by investigator: ___________________________** **Date:** _____ /_______ /_201___

*Please sign*

Brands to be used for nut paste challenges

PEANUT

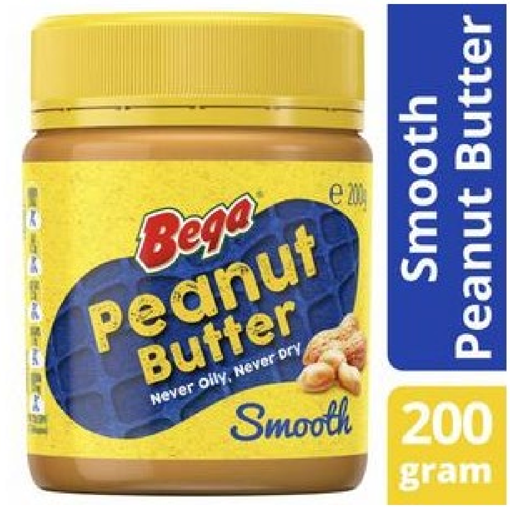

**Bega Peanut Butter Smooth**

**$3.00**

200g | $1.50 per 100G

Add 1 for $3.00

Made in Australia from at least 10% Australian ingredients

Smooth"

Made in Port Melbourne for over 55 years, we use the same recipe t
roasted peanut taste Australians love.

CASHEW

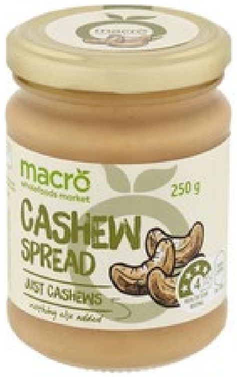

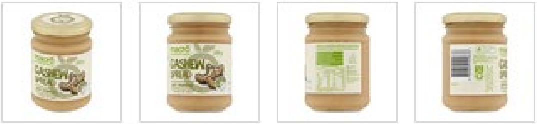

Macro Natural Cashew Spread 250g

$8.00

$3.20 / 100G

Save to list +

Add to cart

Check stock in our stores >

**STANDARD OPERATING PROCEDURE**

**Oral Food Challenge – The OPTIMUM Study**

| | Name | Signature | Date |
|---|---|---|---|
| Author or Updated by | Marie Estcourt | | |
| Reviewed by | Glady Perez | | |
| Approved by | Dr Peter Richmond | | |
| Approved by | Dr Michael O'Sullivan | | |
| Implementation Date | | Insert date | |
| Date to be reviewed (at least annually) | Name | Signature | Date |
| Insert date | | | |

#### 1. Purpose

1. To detail the procedures involved in carrying out the Oral Food Challenge (OFC) Appointment for participants of the OPTIMUM study.
2. To ensure study data from this visit will be accurately and consistently recorded and that the oral food challenge is undertaken in keeping with the policies and procedures of the Immunology Department of Perth Children's Hospital and the OPTIMUM protocol.

#### 2. Scope

1. To provide a procedure for Immunology Research staff to undertake the OFC appointment visit for the OPTIMUM clinical trial

#### 2. Applicable to:

- All investigators, research nurses and research staff involved in performing oral food challenges for the OPTIMUM clinical trial.

#### 4. Definitions and Abbreviations

SOP: Standard Operating Procedure.

SPT: Skin Prick Testing.

OFC: Oral Food Challenge

PCH: Perth Children's Hospital

#### 5. Foods for challenge

Brands to be used for nut paste challenges
<u>PEANUT</u>

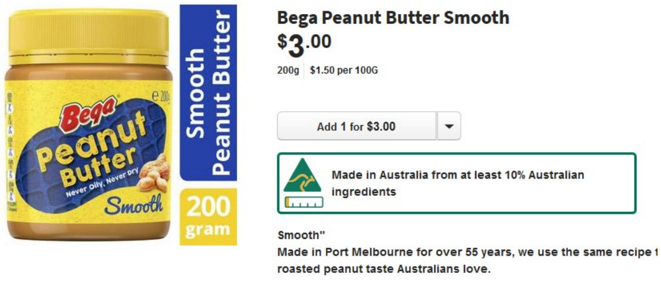

<u>CASHEW</u>

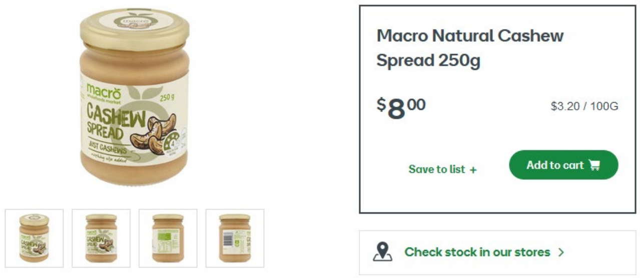

Egg: To be prepared by participant parent according to ASCIA cooking guidelines (below) and measured out by staff;

Preparation instructions

1 Beat one large egg (yolk and white) with a fork for 1 minute

2 Place in microwave

3 Cook at 10 second intervals, mashing the egg mixture with a fork each time.

4 Cook egg until it is rendered semi-solid (approximately a total of 30 seconds of cooking is sufficient)

Milk:To be brought in by participant parent, full cream cows milk.

#### 6. Procedure

Considerations

- It is better to do appointments in the morning to allow for extended observation time if required, without necessitating hospital admission
- Consider any clinical symptoms that may require deferral of the Food Challenge (e.g. current viral illness, poorly controlled asthma or allergic rhinitis) - seek advice from Site Investigator or Delegate prior to administering the first dose.
- Ensure the study participant does not take any form of *Antihistamine (remember to tell parents what drugs constitute antihistamines)* for four days prior to the challenge. If they arrive for OFC having taken antihistamine in the previous four days, the appointment will have to be rescheduled.

**All staff to be familiar with the MET criteria, and recognise when to initiate a MET call (at PCH make the ICU team aware if we give 2 or more doses of IM adrenaline, even if MET team not immediately required).**

##### 5.1 Materials

To be collected from VTG office:

1. Collect participant's folder including CRF from the VTG office at PCH.
    a. Ensure folder includes CRF, OPTIMUM allergy outcome form and food exposure questionnaire(stage 1 participants only).
2. Project clinic container requiring items needed for clinic (peanut butter, etc)

To be collected from Clinical Trials Pharmacy in Research Clinic D on Level 1:

1. OPTIMUM Clinic emergency medication box (containing EpiPens, adrenaline and antihistamine)

##### 5.2 Clinic procedure

***Conducting appointment visit:***
***Requirements:***

- Study doctor and study nurse to be available at all times.
- Resuscitation facilities available.
- Medication doses (including adrenaline) calculated and prescribed on med chart.

***Beginning of the day:***
0830 hours Admit study participant via hospital admitting point (if they are to be admitted)

***Perform Baseline Assessments:***

- Baseline temperature, heart rate, respiratory rate, blood pressure (more frequently if indicated during procedure), oxygen saturations, chest auscultation, skin observations (including SCORAD) and weight. Document results on OFC CRF.
- Weight to calculate Emergency medication doses. Measure height.
- Subject must be haemodynamically stable with observations within satisfactory limits for age; child must weigh more than 7 kg and be afebrile – seek advice from Site Investigator if temperature >38 degrees C.

*NOTE: If the study participant does not satisfy these requirements refer to Site Investigator or delegate for advice.*

***Emergency equipment and resuscitation drugs to be available, doses and volumes appropriate to the study participants weight and age calculated and documented on study form prior to OFC. Draw-up Adrenaline, appropriate for the participant's age and weight.***

**During Food Challenge**

- Challenge food can be mixed with other non-allergenic food upon clinical discretion.
- Ask parent if they have brought in foods to be mixed with challenge food.
- Follow the introduction steps in CRF separated by 20 minutes
- Doses can be repeated upon clinical discretion
- Heart rate, respiratory rate, oxygen saturations, chest auscultation and skin observations every 15 minutes while having OFC and then observe for 2 hours post final dose.
- Record any signs/symptoms of allergic reaction including gastrointestinal, respiratory, cardiovascular, and cutaneous manifestations. Note behaviour changes in younger children.
- Any changes in these parameters of clinical significance should be reported to the Site Investigator or Delegate and managed as per study protocol.
- Observe for 2 hours post challenge
- Stop Food Challenge if any objective signs of an allergic reaction develop (refer to study protocol and Cessation criteria as detailed on CRF).
- Objective symptoms that stop a challenge should be treated as a safety precaution.
- Any treatment or medication will be documented in drug section of OFC CRF and in study participant's progress notes.
- If signs of anaphylaxis develop (e.g .wheeze, stridor, hoarse voice, difficulty breathing, drowsiness, pale/floppy, persistent hypotension), alert the study doctor and administer adrenaline.
- Complete the participant's CRF during the food challenge. Make sure data is clear and accurate enough to enter directly into the database.

***Prior to discharge***

- The participant must wait for 2 hours after completing challenge before leaving or until stable and suitable for discharge. Investigator or delegate to review participant if an allergic reaction has occurred.
- The participant must receive a discharge letter with an ASCIA Action Plan on the back prior to discharge.
- Give family advice based on the outcome
    a. Passed – continue regular food of interest consumption at home (2 tsp, 3 x per week)
    b. Failed – avoid food of interest and food containing food of interest.
- Notify child's GP of outcome of food challenge

##### 5.3 Appointment Completion Procedure

- Ensure participant enrolment details have not changed. If they have changed, update in CRF.
- Thank participant for their continued involvement in the OPTIMUM project and for attending the appointment. Remind participant's parent/caregiver they will be contacted 24 hours after challenge and if they have not attended the 18 and 19 month visits yet they will be booked when the participant is old enough.
- Give participant a discharge letter
- Arrange referral if required (if reacted)
- Check CRF documentation is all completed, if you have missed any details on any questions, telephone the participant the next working day in order to ensure all questions are fully completed. Sign and date name once section is complete.
- If child reacted to food of interest please complete OPTIMUM Allergy Outcome Form
- Take participant's files to the VTG office.
- Return

**7. Amendment History**

| **SOP Version Number** | **SOP Implementation Date** | **Description of Changes** |
|---|---|---|
| 1 | 14/5/2020 | |
