## Supplementary material 2 for "Efficacy of an early first dose of whole-cell pertussis vaccine in infants for the prevention of IgE-mediated food allergy: A randomised controlled trial"

2026-11-06

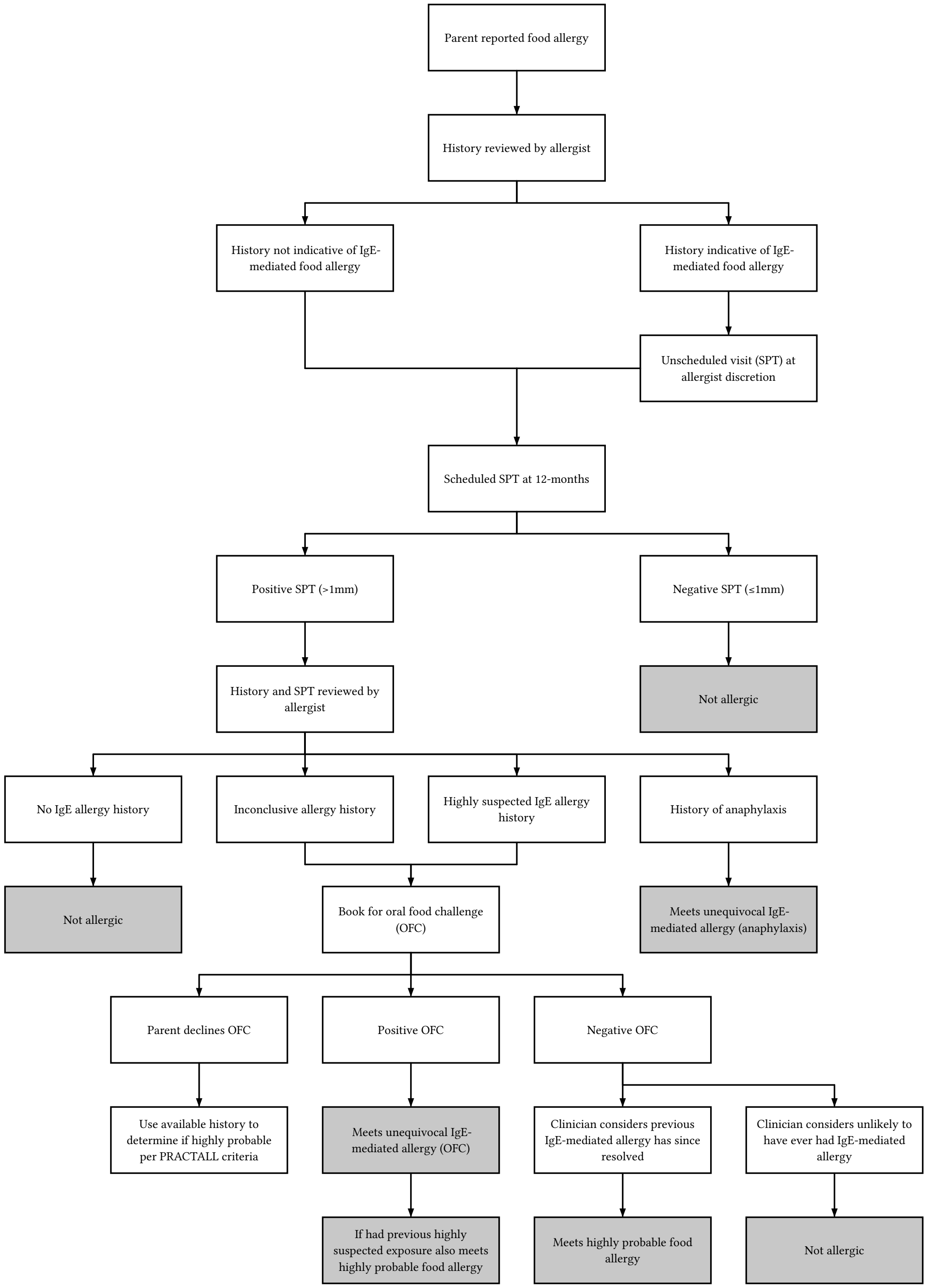


**Figure E1:** Flowchart for determination of allergy outcome.

**Table E1:** Vaccination schedule (4-month/6-month vaccines) received by trial participants stratified by assigned treatment group, '-' indicates unknown vaccination type (or status).

| **Vaccine schedule** | **aP**<br>N = 486 | **wP**<br>N = 486 |
|---|---|---|
| aP3/aP3 | 376 (77%) | 360 (74%) |
| -/aP3 | 33 (6.8%) | 44 (9.1%) |
| aP5/aP5 | 17 (3.5%) | 27 (5.6%) |
| aP3/aP5 | 21 (4.3%) | 16 (3.3%) |
| aP3/- | 15 (3.1%) | 18 (3.7%) |
| aP5/aP3 | 12 (2.5%) | 11 (2.3%) |
| -/- | 5 (1.0%) | 8 (1.6%) |
| -/aP5 | 4 (0.8%) | 1 (0.2%) |
| aP5/- | 3 (0.6%) | 1 (0.2%) |

**Table E2:** Number and percentage of infants who currently eat egg, cow's milk, and peanut by study group via study food and household questionnaire by survey age.

| Food and survey age | **aP** N = 486 | **wP** N = 486 |
|---|---|---|
| Egg | | |
| 6-month | 166/476 (35) | 159/480 (33) |
| 9-month | 406/474 (86) | 409/480 (85) |
| 12-month | 436/477 (91) | 430/478 (90) |
| 15-month | 427/454 (94) | 446/467 (96) |
| 18-month | 447/471 (95) | 441/472 (93) |
| Milk | | |
| 6-month | 135/476 (28) | 139/480 (29) |
| 9-month | 319/474 (67) | 328/480 (68) |
| 12-month | 400/477 (84) | 392/478 (82) |
| 15-month | 412/454 (91) | 423/467 (91) |
| 18-month | 434/471 (92) | 430/472 (91) |
| Peanut | | |
| 6-month | 141/476 (30) | 132/480 (28) |
| 9-month | 397/474 (84) | 395/480 (82) |
| 12-month | 435/477 (91) | 430/478 (90) |
| 15-month | 439/454 (97) | 447/467 (96) |
| 18-month | 452/471 (96) | 447/472 (95) |

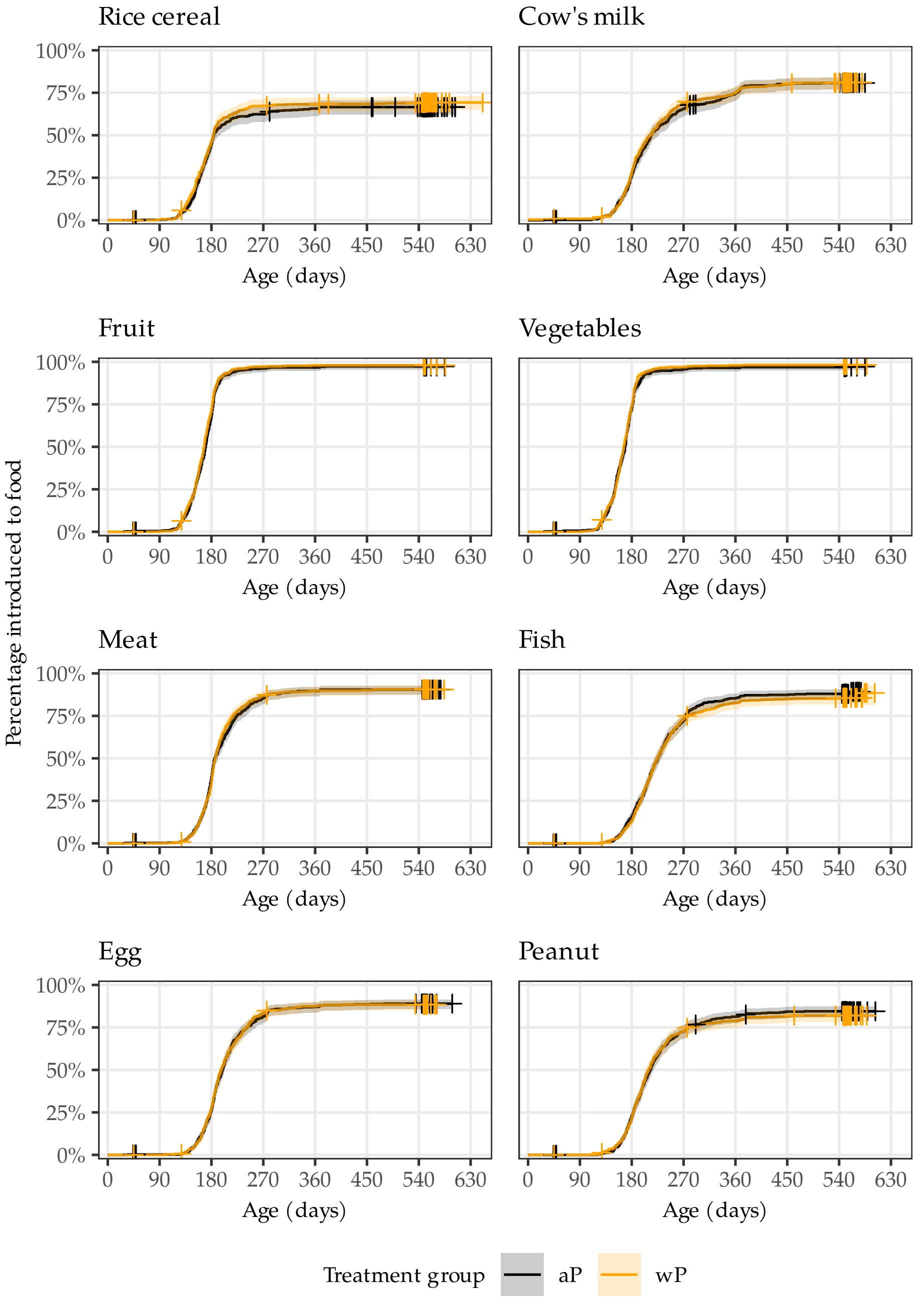


**Figure E2:** Age (days) at food introduction by type of food and assigned study group. Censored observations are marked by a cross.

**Table E3:** Number and proportion with IgE-mediated food allergy and posterior summaries for risk and difference in risk by assigned study group.

| | aP | | wP | | Difference |
|---|---|---|---|---|---|
| | y/n (%) | Median (95% CrI) | y/n (%) | Median (95% CrI) | Median (95% CrI), Pr(<0) |
| Unadjusted | 26/486 (5.3%) | 5.5 (3.7, 7.7) | 31/486 (6.4%) | 6.5 (4.5, 8.9) | 1.0 (-2.0, 4.0), 0.25 |
| Adjusted | 26/486 (5.3%) | 5.4 (3.7, 7.6) | 31/486 (6.4%) | 6.4 (4.5, 8.7) | 1.0 (-1.9, 3.9), 0.26 |

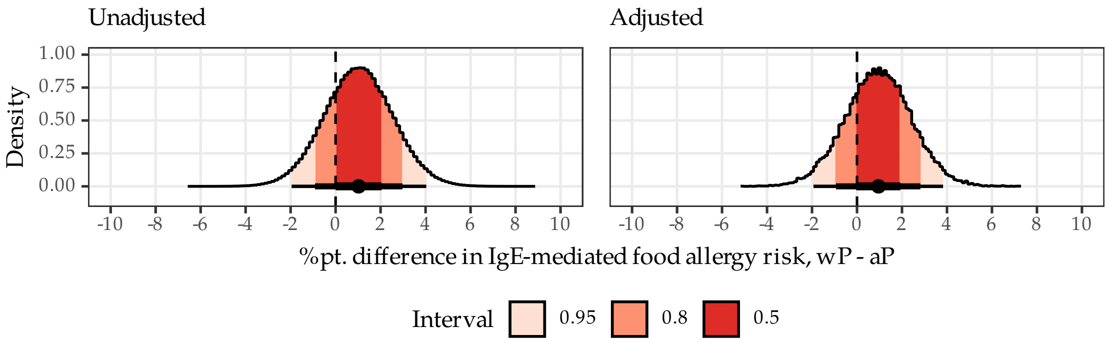


**Figure E3:** Posterior density for difference in risk of IgE-mediated food allergy by model. Unadjusted model was the primary Beta-Binomial model and adjusted model was logistic regression with terms for the specified baseline factors. For the adjusted model, the posterior risk difference was standardised to the trial sample.

| Subgroup | aP | wP | Median (95% CrI) |
|---|---|---|---|
| All participants | 26/486 (5%) | 31/486 (6%) | 1.0 (-2.0, 4.0) |
| Site | | | |
| MCRI | 16/220 (7%) | 14/219 (6%) | -0.9 (-5.6, 3.8) |
| PCH | 6/206 (3%) | 13/207 (6%) | 3.3 (-0.5, 7.5) |
| SCH | 1/30 (3%) | 2/31 (6%) | 2.8 (-7.2, 14.7) |
| CHW | 3/30 (10%) | 2/29 (7%) | -2.8 (-17.7, 10.9) |
| Season of birth | | | |
| Autumn | 8/133 (6%) | 12/138 (9%) | 2.7 (-3.5, 9.0) |
| Winter | 13/144 (9%) | 11/144 (8%) | -1.4 (-7.9, 5.1) |
| Spring | 3/135 (2%) | 6/133 (5%) | 2.2 (-2.0, 6.9) |
| Summer | 2/74 (3%) | 2/71 (3%) | 0.1 (-5.5, 5.8) |
| Family history of atopy | | | |
| No | 2/68 (3%) | 6/71 (8%) | 5.4 (-1.8, 13.6) |
| Yes | 24/418 (6%) | 25/415 (6%) | 0.3 (-2.9, 3.5) |
| Family history of food allergy | | | |
| No | 13/365 (4%) | 19/364 (5%) | 1.6 (-1.2, 4.6) |
| Yes | 13/121 (11%) | 12/122 (10%) | -0.9 (-8.7, 6.8) |
| One parent born in East Asia | | | |
| No | 21/406 (5%) | 20/389 (5%) | -0.0 (-3.1, 3.0) |
| Yes | 4/36 (11%) | 10/48 (21%) | 9.7 (-5.9, 25.0) |
| Unknown | 1/44 (2%) | 1/49 (2%) | -0.2 (-6.7, 6.0) |

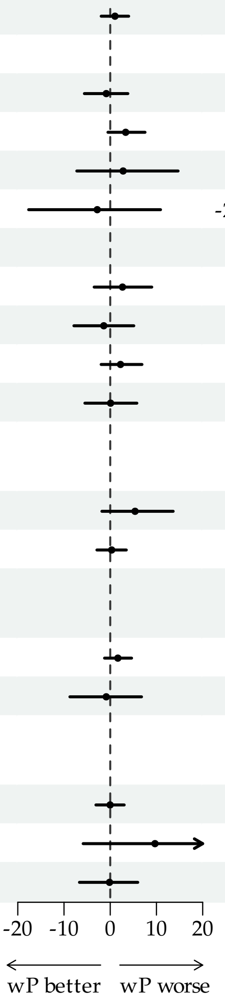


**Figure E4:** Post-hoc subgroup analyses were conducted to explore the effect of group assignment on the primary outcome by season of birth, East Asian birth parent per standard classification, (*Standard Australian Classification of Countries (SACC), 2016 | Australian Bureau of Statistics* 2025) parent-reported history of atopy in a first-degree relative, parent-reported history of IgE-mediated food allergy in a first-degree relative, and site of enrolment. The figures hows subgroup specific estimates of difference in risk (percentage point) of food allergy, wP versus aP. Estimates were derived from logistic regression models with terms for treatment, grouping factor, and their interaction. Diffuse priors were used on all terms.

**Table E4:** Posterior summaries for risk and difference in risk of IgE-mediated food allergy amongst participants with an SPT result.

| | aP | | wP | | Difference |
|---|---|---|---|---|---|
| | y/n (%) | Median (95% CrI) | y/n (%) | Median (95% CrI) | Median (95% CrI), Pr(<0) |
| Unadjusted | 26/473 (5.5%) | 5.6 (3.8, 7.9) | 31/471 (6.6%) | 6.7 (4.7, 9.2) | 1.1 (-2.0, 4.2), 0.24 |
| Adjusted | 26/473 (5.5%) | 5.6 (3.7, 7.9) | 31/471 (6.6%) | 6.6 (4.6, 9.0) | 1.0 (-2.0, 4.0), 0.26 |

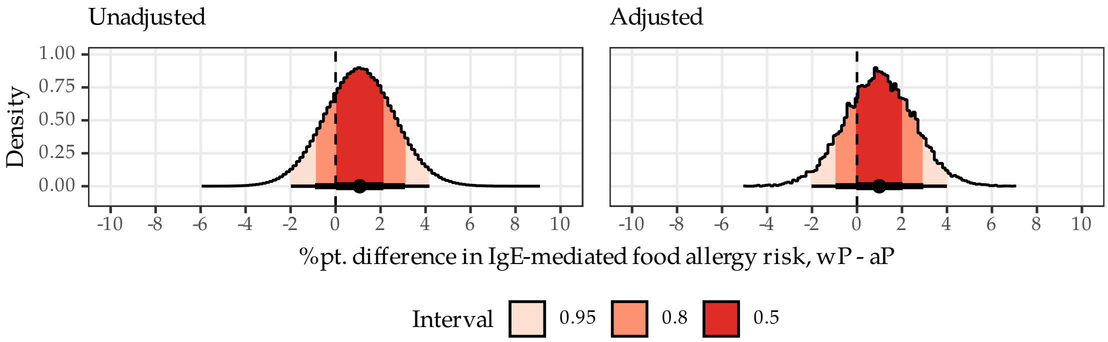


**Figure E5:** Posterior density for difference in risk of IgE-mediated food allergy by model amongst participants with an SPT result. Unadjusted model was the primary Beta-Binomial model and adjusted model was logistic regression with terms for the specified baseline factors. For the adjusted model, the posterior risk difference was standardised to the trial sample.

**Table E5:** Posterior summaries for risk and difference in risk of eczema by age of onset.

| | aP | | wP | | Difference |
|---|---|---|---|---|---|
| | y/n (%) | Median (95% CrI) | y/n (%) | Median (95% CrI) | Median (95% CrI), Pr(<0) |
| Unadjusted | | | | | |
| 6m | 34/483 (7.0%) | 7.2 (5.1, 9.7) | 28/478 (5.9%) | 6.0 (4.1, 8.3) | -1.2 (-4.3, 2.0), 0.77 |
| 12m | 47/482 (9.8%) | 9.9 (7.4, 12.7) | 43/478 (9.0%) | 9.1 (6.8, 11.9) | -0.7 (-4.5, 3.0), 0.65 |
| Adjusted | | | | | |
| 6m | 34/483 (7.0%) | 7.2 (5.1, 9.6) | 28/478 (5.9%) | 5.8 (4.0, 8.1) | -1.3 (-4.3, 1.7), 0.80 |
| 12m | 47/482 (9.8%) | 9.8 (7.4, 12.7) | 43/478 (9.0%) | 9.0 (6.7, 11.7) | -0.8 (-4.5, 2.7), 0.68 |

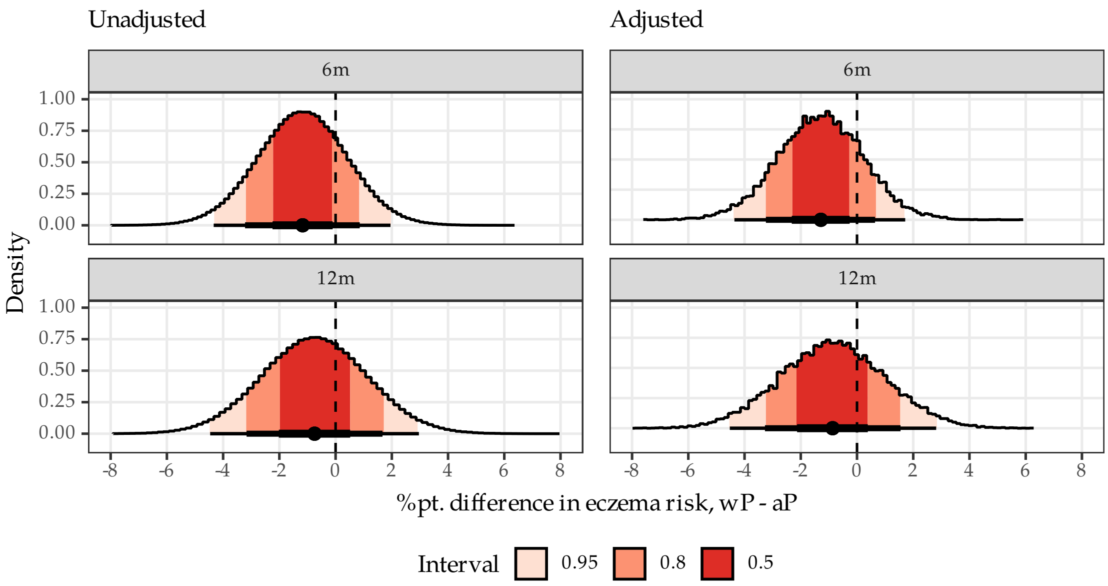


**Figure E6:** Posterior density for difference in risk of new-onset eczema by model. For the adjusted model, the risk difference was standardised to the trial sample.

**Table E6:** Posterior summaries for risk and difference in risk of positive skin prick test by size.

| | aP | | wP | | Difference |
|---|---|---|---|---|---|
| | y/n (%) | Median (95% CrI) | y/n (%) | Median (95% CrI) | Median (95% CrI), Pr(<0) |
| Unadjusted | | | | | |
| >1mm vs ctrl. | 81/473 (17%) | 17.2 (14.0, 20.8) | 86/471 (18%) | 18.4 (15.0, 22.0) | 1.1 (-3.7, 6.0), 0.32 |
| ≥3mm vs ctrl. | 58/473 (12%) | 12.4 (9.6, 15.5) | 61/471 (13%) | 13.1 (10.2, 16.3) | 0.7 (-3.6, 4.9), 0.38 |
| Adjusted | | | | | |
| >1mm vs ctrl. | 81/473 (17%) | 17.3 (14.0, 20.8) | 86/471 (18%) | 18.2 (15.0, 21.7) | 1.0 (-3.8, 5.6), 0.35 |
| ≥3mm vs ctrl. | 58/473 (12%) | 12.4 (9.7, 15.5) | 61/471 (13%) | 12.9 (10.1, 16.1) | 0.5 (-3.7, 4.7), 0.40 |

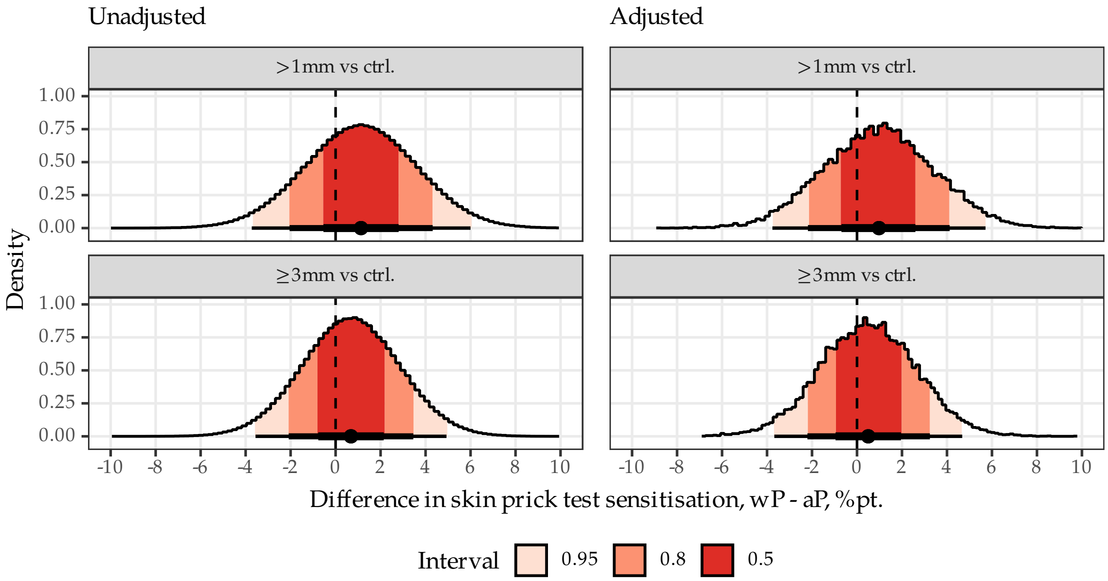


**Figure E7:** Posterior density for difference in risk of skin prick test sensitisation by model. For the adjusted model, the risk difference was standardised to the trial sample.

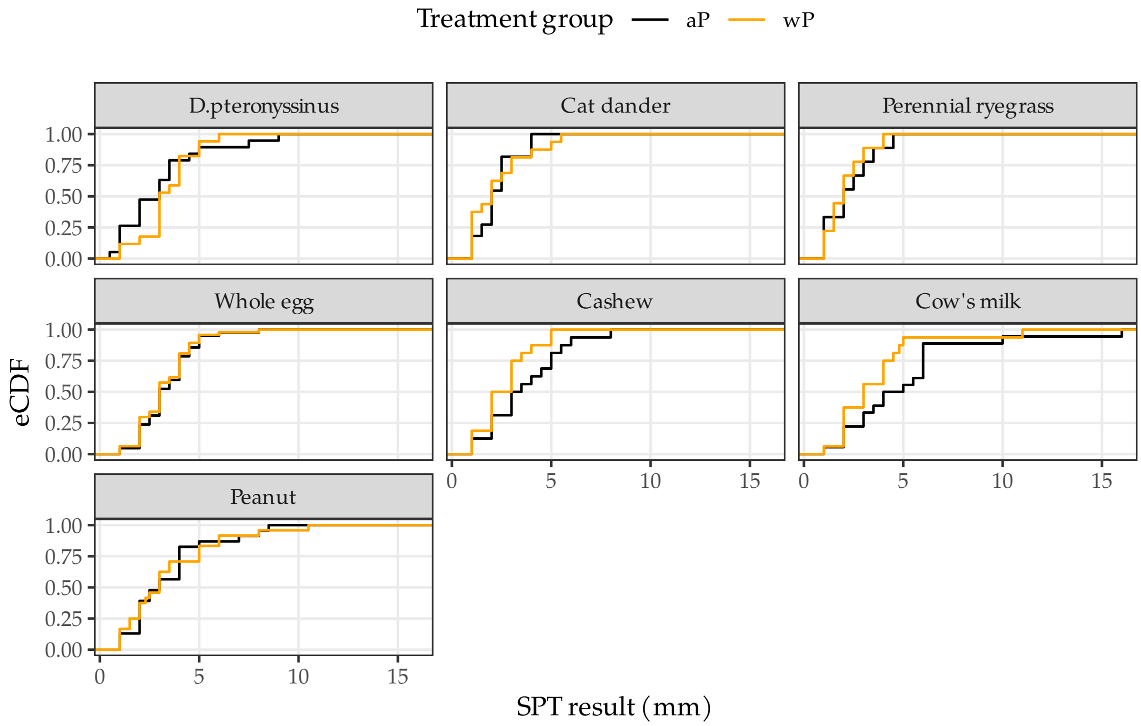


**Figure E8:** Wheal diameter size amongst participants with an SPT reaction (diameter >0mm) by tested allergen and assigned study group.

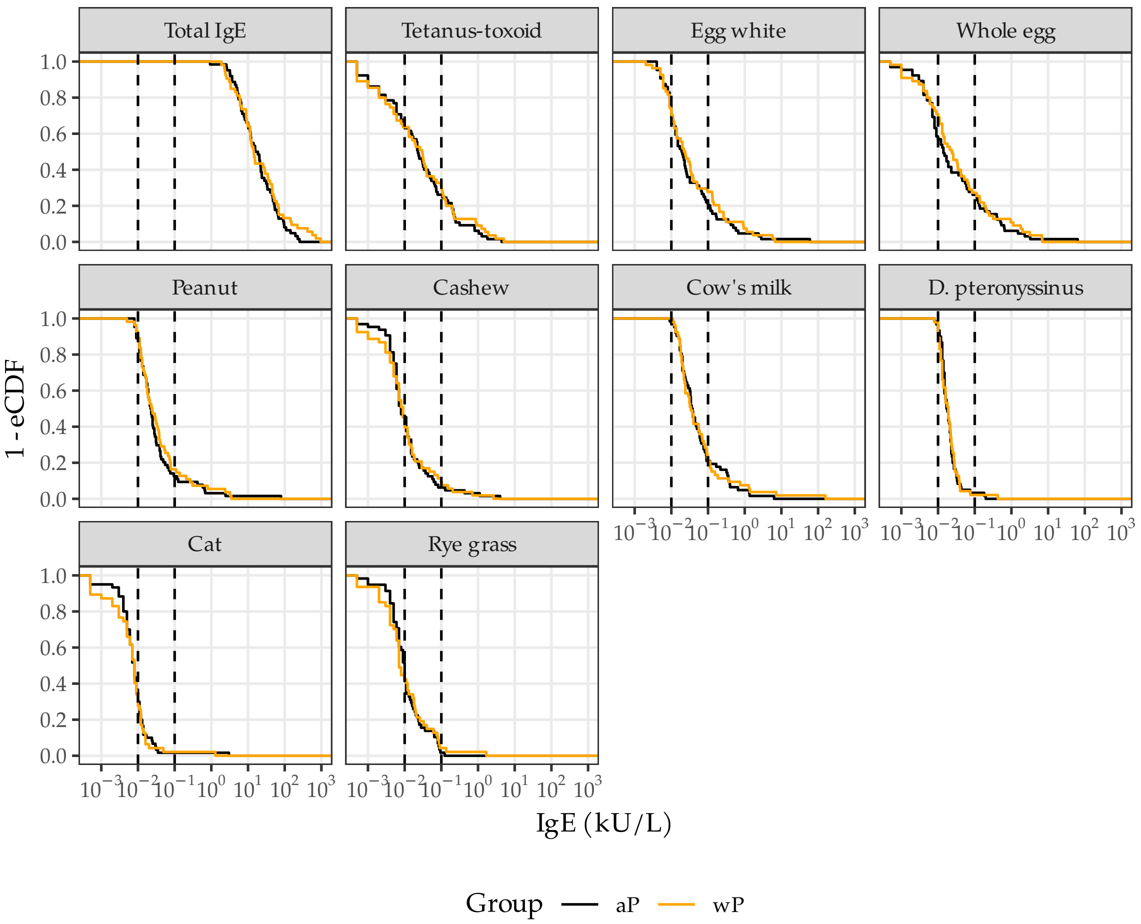


**Figure E9:** Distribution of total and allergen-specific IgE concentrations (kU/L) by assigned study group. Vertical dotted lines indicate 0.01 and 0.1 kU/L

**Table E7:** Listing of serious adverse events.

| Participant SAE ID | Treatment | MedDRA code | Severity | Type of medical advice sought | Relationship to study vaccine? | Study day |
|---|---|---|---|---|---|---|
| 1 | wP | Bronchiolitis | Mild | Hospitalisation | Not Related | 144 |
| 2 | wP | Escherichia urinary tract infection | Moderate | Hospitalisation | Not Related | 43 |
| 2 | wP | Hospitalisation | Mild | Hospitalisation | Not Related | 95 |
| 2 | wP | Investigation | Mild | Hospitalisation | Not Related | 190 |
| 3 | wP | Bronchiolitis | Moderate | Hospitalisation | Not Related | 49 |
| 4 | aP | Hospitalisation | Mild | Hospitalisation | Not Related | 7 |
| 5 | wP | Rectal haemorrhage | Moderate | Hospitalisation | Not Related | 7 |
| 6 | aP | Ileocolic intussusception | Severe | Hospitalisation | Not Related | 189 |
| 7 | aP | Laryngomalacia | Mild | Hospitalisation | Not Related | 48 |
| 8 | aP | Urinary tract infection | Moderate | Hospitalisation | Not Related | 46 |
| 9 | aP | Gastroenteritis NOS | Moderate | Hospitalisation | Not Related | 19 |
| 10 | wP | Irritability postvaccinal | Moderate | Hospitalisation | Related | 0 |
| 11 | wP | Bronchiolitis | Moderate | Hospitalisation | Not Related | 191 |
| 11 | wP | Bronchiolitis | Moderate | Hospitalisation | Not Related | 209 |
| 12 | wP | Benign rolandic epilepsy | Moderate | Hospitalisation | Not Related | 154 |
| 13 | wP | Gastrooesophageal reflux | Severe | Hospitalisation | Not Related | 3 |
| 14 | aP | Herniotomy | Moderate | Hospitalisation | Not Related | 29 |
| 15 | wP | Acute bronchiolitis | Severe | Hospitalisation | Not Related | 42 |

### References

*Standard Australian Classification of Countries (SACC), 2016 | Australian Bureau of Statistics*. 2025. https://www.abs.gov.au/statistics/classifications/standard-australian-classification-countries-sacc/latest-release.
